# EBV deletions as biomarkers of response to treatment of Chronic Active Epstein Barr Virus

**DOI:** 10.1101/2020.12.18.20248315

**Authors:** Cristina Venturini, Charlotte J Houldcroft, Arina Lazareva, Fanny Wegner, Sofia Morfopoulou, Persis J. Amrolia, Zainab Golwala, Anupama Rao, Stephen D. Marks, Jacob Simmonds, Tetsushi Yoshikawa, Paul J. Farrell, Jeffrey I. Cohen, Austen J. Worth, Judith Breuer

## Abstract

Chronic active Epstein Barr Virus (CAEBV) is a rare condition occurring in previously healthy individuals associated with persistent EBV viraemia, fever, lymphadenopathy and hepatosplenomegaly. Viral deletions have been found in CAEBV and other lymphomas. However, it is unclear how stable these deletions are, whether they are present in different sites and how they evolve overtime. We sequenced fourteen longitudinal blood samples from three European CAEBV patients and compared with CAEBV saliva samples and other sequences from EBV-related conditions. We observed large EBV deletions in blood, but not saliva from CAEBV patients. Deletions were stable over time but were lost following successful treatment. Our results are consistent with the likelihood that certain deletions in the virus from CAEBV patients are associated with the evolution and persistence of haematological clones. We propose that the loss of deletions following successful treatment should be investigated as a potential biomarker to aid CAEBV management.

## Introduction

Epstein-Barr virus (EBV) infects >95% of the population worldwide (Longnecker et al., 2013). A small number of patients develop life-threatening persistence of high level/sustained EBV replication following an infectious mononucleosis syndrome, often associated with splenomegaly and hepatitis (Cohen et al., 2011). Chronic active EBV (CAEBV) (see “Clinical definitions” in Supplementary materials) is characterised by infiltration of tissues by EBV positive T, NK or less frequently B cells and can progress into lymphoproliferative disease. Clonal expansion of EBV-infected T or NK cells containing identical EBV genome and chromosomal abnormalities is well described with both some oligo and polyclonal populations of EBV genomes found (Kimura, 2006; Ohshima et al., 1997; Quintanilla-Martinez et al., 2000).

To date, CAEBV has been mostly described in Asian or South/Central American patients (Kimura & Cohen, 2017). Frequent deletions in the EBV genome have been found in samples from Japanese CAEBV patients (35%) and other EBV-driven neoplasms (Murata et al., 2020; Okuno et al., 2019; Peng et al., 2019). However, it is unclear if these deletions are specific for Asian EBV strains, whether they are present in viral genomes in different sites and how they evolve overtime.

We sequenced EBV from serial blood samples obtained from three UK patients with CAEBV. The results were compared with sequences from saliva of CAEBV patients and blood and tissue from other benign and malignant EBV-related conditions.

## Results and discussion

### EBV genomes in blood and saliva from CAEBV patients

EBV blood genomes from CAEBV patients clustered with European/US sequences; longitudinal samples, including those after PBSCT, clustered by patient (Supplementary figure 2). Within-host CAEBV nucleotide diversity (π) was low and comparable to that of EBV from other blood samples (Table S1 and Supplementary figure 3-4). In contrast CAEBV salivary samples showed significantly higher EBV diversity compared to blood and tumour samples (p<0.001), indicating the presence of multiple strains (Supplementary figure 5-7). Single nucleotide variants (SNVs) and small (<2kbp) deletions were present in all samples, largely located in latent genes as well as BPLF1 and BLLF1 [gp350] (Supplementary figures 8-16). Our results showed that EBV shed in saliva has the hallmarks of lytic replication and frequent mixed infections in line with previous studies (Kwok et al., 2015; Renzette et al., 2014).

### Larger Deletions

We identified low frequency larger (>2kb) EBV deletion in patient 1, as well as PTLD (2/4) and HL (2/7) tumour-tissue at position 120470-158062 (Figure 1). This includes the BART miRNA clusters, as well as several lytic genes, including scaffold proteins (i.e. BdRF1, BVRF2), glycoproteins (i.e. BILF2, BXLF2), a tegument protein (BVRF1), and regulators of late gene transcription (BcRF1, BVLF1). Interestingly, patient 1 also had a nonsense mutation in BXLF2 (Supplementary Figure 17). A second larger deletion at positions 12118-15159, which is part of the major EBV repeat (IR1) was present in blood from CAEBV patients (4/4) and one patient with PID.

**Figure 1A:**
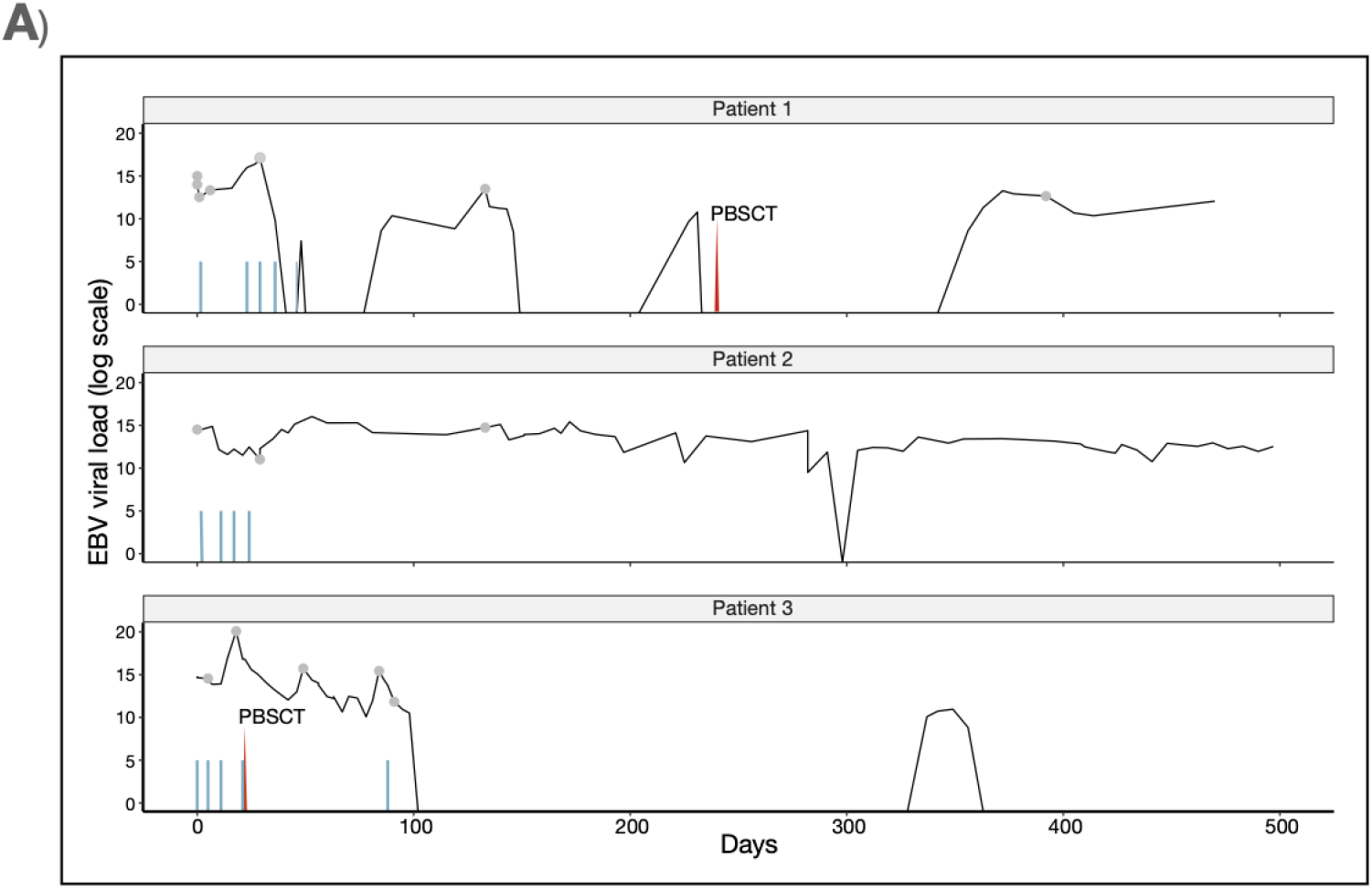
Viraemia and treatment in GOSH CAEBV patient. Patient 1 and 3 received PBSCT (in red) and all patients received 4 to 5 doses of rituximab (blue lines). Samples sequenced are indicated in grey filled dots.

**Figure 1B:**
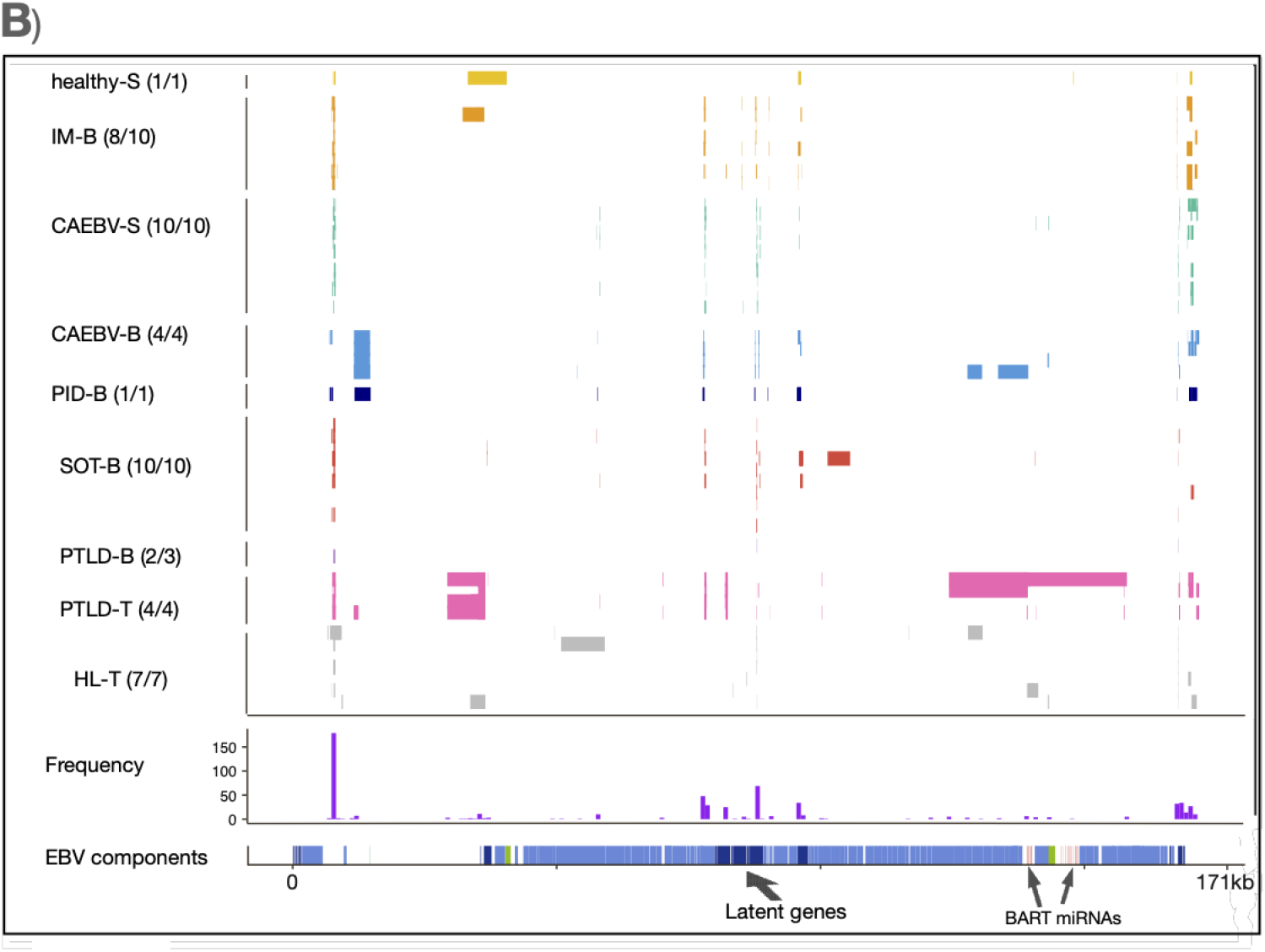
Summary of deletions (≥ 30 bp). Samples for healthy (healthy-S), infectious mononucleosis (IM-B), CAEBV (CAEBV-S, CAEBV-B), primary immunodeficiency (PID) as well as EBV-positive solid organ transplant (SOT-B), PTLD (PTLD-B and PTLD-T) and HL (HL-T). Each line represents an EBV genome from a single patient (if multiple samples from one patient were available, only the one with the highest read depth was included here).

**Figure 1C:**
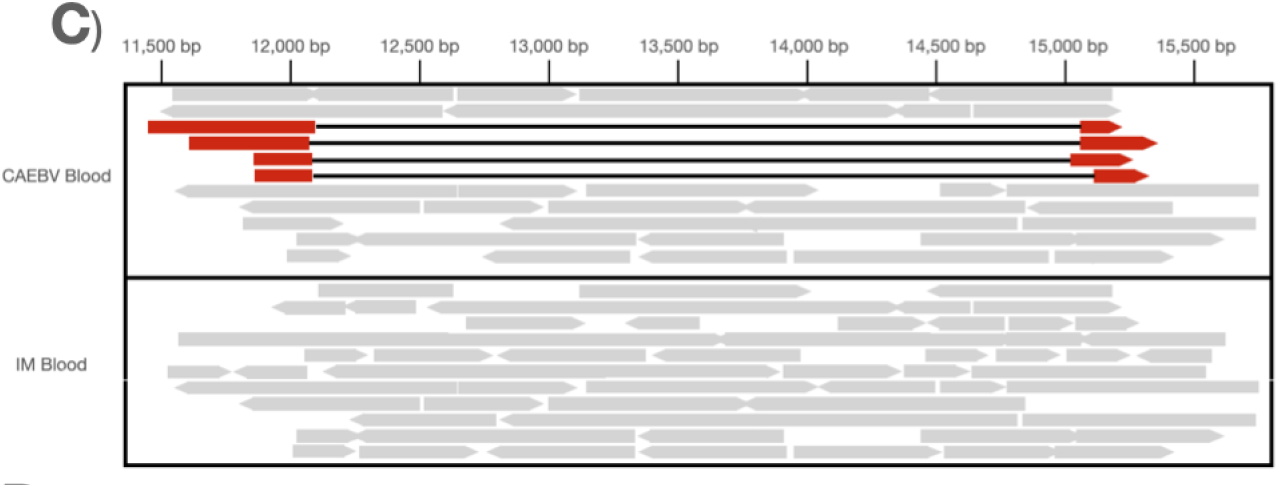
Representation of the IR1 deletion in CAEBV. Representative portion of the alignment file for a blood CAEBV sample to demonstrate IR1 deletion (represented by red reads and black lines) compared to an IM blood sample where the deletion is not present (reads in grey).

**Figure 1D:**
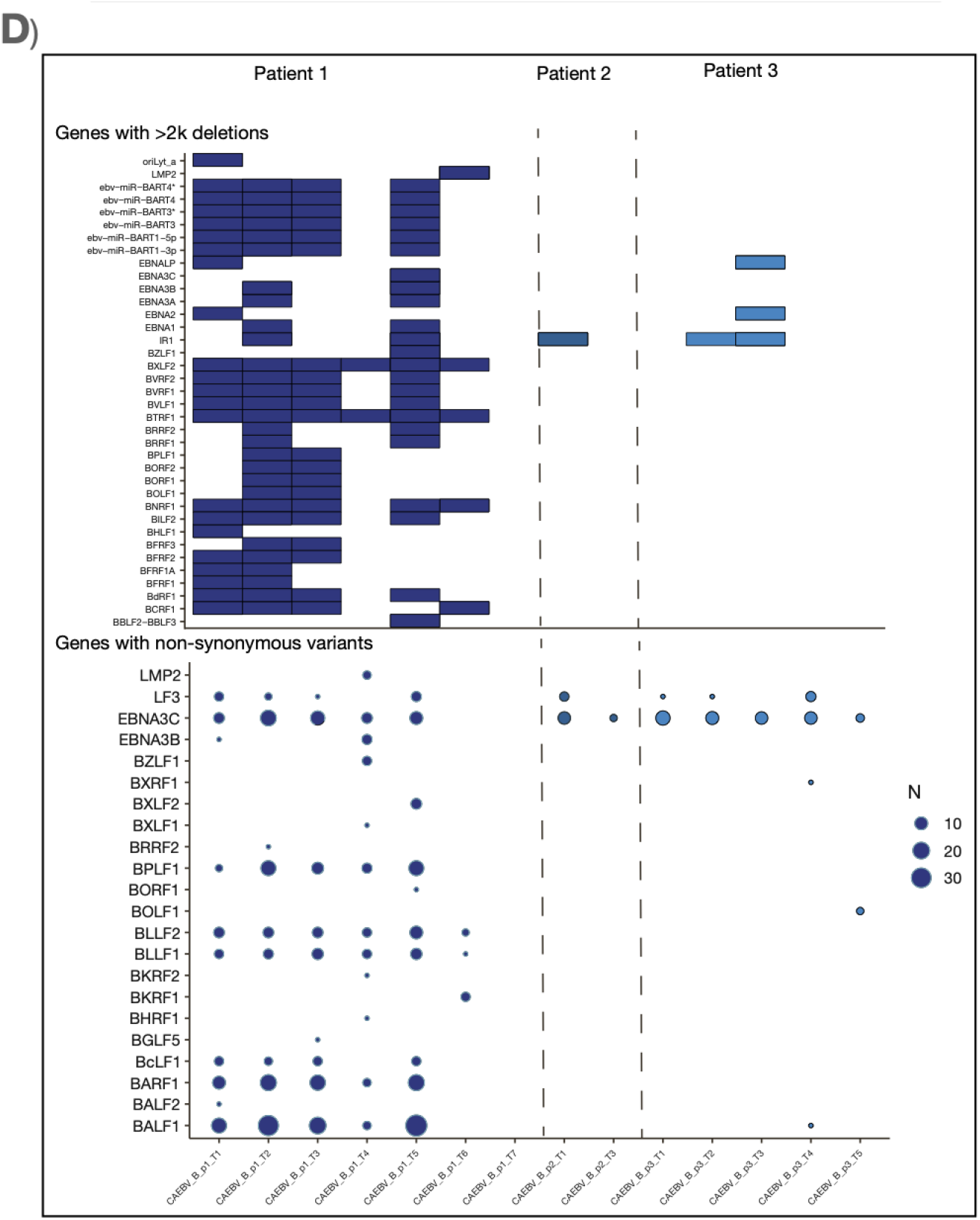
Analysis of longitudinal samples in CAEBV patients. Genes affected by larger deletions (≥ 2k) are shown in the heatmap; overtime all larger deletions are lost. Non-synonymous low frequency variants (<50%) are shown in the bubble plot.

### EBV variation overtime in CAEBV patients

Analysis of variation overtime revealed that the blood from two CAEBV patients (1 and 3) with EBV present predominantly in T cells, showed higher EBV genomic heterogeneity with increasing number of low frequency (<50%) variants and deletions than patients with IM (Figure 1D, Supplementary figures 18-19). In contrast, patient 2, whose EBV was only found in B cells, had fewer SNVs and deletions with a picture similar to that seen in the blood from patients with PTLD with evidence of clonality. In patient 2, larger deletions were stable overtime but were lost after rituximab (Figure 1D). In CAEBV patient 1, larger deletions persisted despite rituximab. Associated clinical deterioration necessitated PBSCT (Table 1), following which both SNVs and deletions were absent from virus reactivating asymptomatically. CAEBV patient 3, who also failed to respond to rituximab, lost most deletions, including the 12118-15159 IR1 deletion following PBSCT.

**Table 1:** Patients descriptions and clinical details

| Pat | Sex | Description | Treatment | Transplant | Reactivation | N samples |
| --- | --- | --- | --- | --- | --- | --- |
| P1 | M | EBV driven HLH<br>primary and secondary immunodeficiency excluded | HLH 94 protocol<br>5 doses of Rituximab (pre-transplant) | Alemtuzumab/Fludarabine/Melphalan conditioned matched unrelated donor (MUD) peripheral blood stem cell transplant (PBSCT) | 4 months post HST had reactivation of EBV without signs of PTLD and resolved without therapy | 7 |
| P2 | F | EBV driven HLH<br>primary and secondary immunodeficiency excluded | HLH 94 protocol<br>4 doses of Rituximab | The patient did not proceed to BMT as she responded well to HLH treatment and had no evidence of ongoing HLH activity despite ongoing EBV viraemia | Remained EBV PCR positive for 3.5 years without symptoms. As of November 2018, EBV PCR is negative. | 3 |
| P3 | F | EBV driven HLH, spontaneously resolved without treatment.<br>Over the following 6 months she had two further EBV-related HLH relapses treated with steroids. Although her HLH associated symptoms rapidly responded, her EBV viraemia continued to rise.<br>primary and secondary immunodeficiency excluded<br>During the 3rd HLH relapse treatment course patient had multiple complications (renal failure requiring haemofiltration, atrial fibrillation requiring DC cardioversion, adenovirus and CMV viremias, invasive candida parapsilosis enteritis, facial palsy) | 4 doses of rituximab with successfully depleted B cells but not reduced EBV viral load (pre-transplant)<br>After 3rd HLH relapse was commenced on etoposide as per HLH 94 protocol with resolution of clinical symptoms and reduction in EBV viral load.<br>1 dose of rituximab after transplant | Alemtuzumab/Fludarabine/Treosulfan conditioned matched sibling donor (MSD) peripheral blood stem cell transplant | EBV reactivation that resolved after 1 dose of Rituximab. | 5 |

### Discussion

EBV deletions were previously described only in CAEBV and malignancies patients of Asian origins (Murata et al., 2020; Okuno et al., 2019; Peng et al., 2019), however we show that these deletions occur independently of geographic origin. Data from mouse models suggest that loss of the BART miRNAs and late lytic genes within 120470-158062 deletion may drive a more lytic phenotype and faster cell growth, this predisposing to tumour formation (Lin et al., 2015; Ma et al., 2011; Murata et al., 2020). Although, the mechanism is not entirely clear, Murata and colleagues argue that the abortive EBV replication resulting from the absence of these regions prevents normal cell death associated with lytic replication (Murata et al., 2014, 2020).

The 12118-15159 deletion overlaps with the IR1 which includes BWRF1 and Wp promoters. There is no direct evidence yet that BWRF1 is biologically significant, however it is known that some components are essential for the transformation of cells by EBV. Our deletion is similar to one identified in strain L591, a Hodgkin’s lymphoma cell line (breakpoint 125bp upstream of hairpin) (Palser et al., 2015). Persistence in longitudinally sampled blood of larger deletions has been associated with clonal expansion of the cells in which they are found (Okuno et al., 2019). Importantly we show that clinical response to therapy is associated with loss of deletions in virus recovered after treatment. Our findings of low-level potentially premalignant clones together with evidence of replicating virus in blood from patients with CAEBV fits with the hypothesis that deletions occur only when a particular subset of the patient’s blood cells are infected with EBV. Progenitor lymphoid cells have been suggested to be the target cell type (Murata et al., 2014).

In conclusion, we confirm that clones containing EBV genomes with specific large deletions are found in blood but not saliva from CAEBV patients. While rituximab reduced the burden of EBV deletions in the blood of B cell-associated CAEBV, it had no impact on the prevalence of EBV deletions in T/NK-associated CAEBV. The absence of these EBV-deletions from virus reactivating following PBSCT suggests the loss of the pre-malignant clone and raises the possibility that these large deletions could be useful biomarkers for monitoring the success of treatments for CAEBV.

## Materials and Methods

### EBV dataset

We analysed a total of 94 EBV genome samples from nine groups: saliva from a healthy asymptomatic EBV carrier (healthy-S), blood from persons with infectious mononucleosis (IM-B), saliva (CAEBV-S) and blood (CAEBV-B) from persons with CAEBV, blood from a patient with persistent EBV viraemia and diagnosis of primary immunodeficiency (LRBA deficiency) (PID-B), blood from asymptomatic patients with EBV following organ transplantation (SOT-B), patients with post-transplant lymphoproliferative disease (PTLD-B) and tumour (PTLD-T) from patients with PTLD, and tumours from patients with Hodgkin lymphoma (HL-T). The samples and patients sequenced, and accession numbers are shown in Supplementary Table1 (Table S1).

### Ethics and sample collection

EBV was sequenced directly from residual diagnostic whole blood samples stored at Great Ormond Street Hospital for Children NHS Foundation Trust (GOSH) at −80 C and obtained by the UCL Infection DNA Bank for use in this study. All samples were supplied to the study in an anonymised form.

Approval for use of anonymized residual diagnostic specimens were obtained through the University College London/University College London Hospitals (UCL/UCLH) Pathogen Biobank National Research Ethics Service Committee London Fulham (Research Ethics Committee reference: 12/LO/1089).

Saliva samples for CAEBV patients in US were approved by the National Institute of Allergy and Infectious Diseases and all subjects provided consent.

### DNA extraction, library construction, targeted enrichment, and sequencing

Total DNA was extracted from each sample using the EZ1 Virus kit and EZ1 XL extraction system (Qiagen) according to manufacturer’s instructions. Virus loads were established by an in-house NHS diagnostic qPCR assay (GOSH).

### SureSelectXT Target Enrichment: RNA baits design

A library of 120-mer RNA baits spanning the length of 124 partial and complete EBV genomes from GenBank were designed using an in-house PERL script. Baits specificity was verified by BLASTn searches against the Human Genomic plus Transcript database. The custom designed EBV baits were uploaded to SureDesign and synthesised by Agilent Technologies.

### SureSelectXT Target Enrichment: Library preparation, hybridisation and enrichment

Total DNA from clinical samples was quantified using the Qubit dsDNA HS assay kit (Life Technologies, Q32854) and between 200-500 ng of DNA was sheared for 150 seconds, using a Covaris E220 focused ultra-sonication system (PIP 175, duty factor 5, cycles per bburst 200). End-repair, non-templated addition of 3’ poly A, adapter ligation, hybridisation, PCR (12 cycles pre-capture and 18 or 22 cycles post capture) and all post-reaction clean-up steps were performed according to the SureSelectXT Automated Target Enrichment for Illumina Paired-End Multiplexed Sequencing 200 ng protocol (version F.2) on the Bravo platform WorkStation B (Agilent Technologies). All recommended quality control steps were performed on the 2200 TapeStation (Agilent Technologies).

### Virus assembly

First, reads were trimmed using TrimGalore (https://www.bioinformatics.babraham.ac.uk/projects/trim_galore/) with a quality threshold of 20 and then aligned with BLASTN against a database of 124 EBV type 1 and 2 genomes in order to extract EBV-specific reads. They were then de novo assembled using SPAdes (Bankevich et al., 2012) into contigs, which were filtered for those of greater length than 200 bp with Quast. The order and orientation of contigs were determined with BLASTN and a scaffold being generated using an in-house R script. The quality checked reads were re-mapped against this pseudogenome using BBMap (https://jgi.doe.gov/data-and-tools/bbtools/bb-tools-user-guide/bbmap-guide/) and further processed into BAM-files with Samtools (v 1.9) (Li et al., 2009) and Picard (https://broadinstitute.github.io/picard/). The consensus sequences were extracted with an in-house PERL script.

Reference-based mapping was also performed for SNVs and deletions detection. The reference used was NC_007605.1 and mapping was performed with Bbmap (https://jgi.doe.gov/data-and-tools/bbtools/bb-tools-user-guide/bbmap-guide/).

### Alignment and phylogenetic analysis

Sequences were aligned with Mafft (v.7) (Katoh et al., 2002) using default parameters. Multiple alignment was then checked manually with Aliview (Larsson, 2014). Distances and clustering were performed with custom R scripts, specifically using the dist.dna function in the “ape” package (Paradis & Schliep, 2019) and cmdscale, which implements classical multidimensional scaling (MDS).

### Diversity calculation and haplotype reconstruction

Diversity calculations have been described elsewhere (Cudini et al., 2019) and code is available here https://github.com/ucl-pathgenomics/NucleotideDiversity. For haplotype reconstruction, we used HaROLD, which uses co-varying variant frequencies in a probabilistic framework. Validation and applications are described here (Cudini et al., 2019; Pang et al., 2020) and the program can be found here https://github.com/ucl-pathgenomics/HaROLD.

### Minority variant analysis

After trimming with TrimGalore, fastq files were mapped to Genbank reference NC_007605.1 using BBmap. Duplicates were removed with Picard and bam files sorted with Samtools. Consensus files were extracted with mpileup (included in Samtools) and QUASR (https://github.com/simonjwatson/QUASR). To focus our analysis only on minority variants (to capture within-host variation and not strain differences with the reference), we took the consensus file and mapped each sample to its own sequence. SNVs were then called with callvariants.sh in the BBmap suite if the frequency was >= 2%, read count >=8, strand ratio of 20% and callvariants.sh quality score > 0.8. SNVs in repetitive regions were excluded according to NC_007605.1 positions. Genes annotation and plots were done using custom R scripts.

### Deletions detection

After trimming with TrimGalore, fastq files were mapped to Genbank reference NC_007605.1 using BBmap allowing for reads to be split across the genome with default parameters. Duplicates were removed with Picard and bam files sorted with Samtools. Deletions were called with callvariants.sh in the BBmap suite with at least 5 reads supporting the split. Gene annotations and plots were done using custom R scripts.

## Data Availability

Sequence reads for CAEBV and PID patients have been deposited in NCBI Sequence Read Archive under BioProject ID PRJEB41945. All accession numbers for the rest of the dataset are available in Table S1.

## Acknowledgements/Funding

We thank Prof. Richard Goldstein (Division of Infection & Immunity, UCL) and Dr Daniel P. Depledge (NYU Department of Medicine) for insightful comments and discussion. We acknowledge the support of the MRC/NIHR funded Pathogen Genomics Unit.

CV was supported by the Wellcome Trust Collaborative Award [204870/Z/16/Z]; SM by a WT Henry Wellcome fellowship [206478/Z/17/Z]. PJF was supported by MRC grant MR/S022597/1 and by NIHR Imperial BRC. JB is supported by the NIHR UCL/UCLH BRC. This work was supported by the intramural research program of the National Institute of Allergy and Infectious Diseases.

## Data availability

Sequence reads for CAEBV and PID patients have been deposited in the European Nucleotide Archive (ENA) under BioProject ID PRJEB41945. All accession numbers for the rest of the dataset are available in Table S1.

## Abbreviations

EBV: Epstein-Barr Virus
CAEBV: Chronic active Epstein Barr Virus
PBSCT: peripheral blood stem cell transplant
HLH: Hemophagocytic Lymphohistiocytosis
IM: Infectious Mononucleosis
PTLD: Post Transfusion Lymphoproliferative Disease
HL: Hodgkin’s lymphoma
PID: Primary Immunodeficiency.

