## Supplementary Material for "EBV deletions as biomarkers of response to treatment of Chronic Active Epstein Barr Virus"

### Clinical definitions

**Chronic active EBV (CAEBV):** CAEBV is defined as a systemic EBV-positive polyclonal, oligoclonal, or (often) monoclonal lymphoproliferative disorder characterized by fever, persistent hepatitis, hepatosplenomegaly, and lymphadenopathy, which shows varying degrees of clinical severity depending on the host immune response and the EBV viral load (WHO definition ^1^). The diagnostic criteria of CAEBV include IM-like symptoms persisting for >3 months, increased EBV DNA (>10^2.5^ copies/μg DNA) in peripheral blood, histological evidence of organ disease, and demonstration of EBV RNA or viral proteins in affected tissues in patients without known immunodeficiency, malignancy, or autoimmune disorders ^1,2^.

### Clinical information

**Patient 1** presented in June 2014 with a high EBV viral load (2.7x10^7^ copies of EBV/mL whole blood) and EBV driven Haemophagocytic lymphohystiocitosis (HLH). Primary and secondary immunodeficiency was excluded, so a diagnosis of T cell CAEBV was considered. The patient was treated with HLH 94 protocol, rituximab (total of 5 x 375mg/m^2^ doses) and cidofovir. After an initial good response, the patient had EBV reactivation after 2 months with HLH symptoms requiring re-initiation of HLH therapy and proceed to Alemtuzumab/Fludarabine/Melphalan conditioned matched unrelated donor (MUD) peripheral blood stem cell transplant (PBSCT) in February 2015. 4 months post HST EBV reactivation was observed, without signs of PTLD and it resolved without therapy. The patient also received Foscarnet for HCMV reactivation post-transplant.

**Patient 2** presented with EBV driven HLH in September 2014; primary and secondary immunodeficiency was excluded, and she was treated as per HLH-94 protocol and Rituximab (4 doses). No genetic cause for HLH was found (on Bridge or WGS). Since she responded well to HLH treatment and had no evidence of ongoing HLH activity despite ongoing EBV viraemia MDT decision not to proceed to BMT. Chronic asymptomatic EBV viraemia persisted for 3.5 years without symptoms. EBV PCR is negative as for November 2018.

**Patient 3** presented with EBV driven HLH in 2014, that spontaneously resolved without treatment. Over the following 6 months she had two further EBV-related HLH relapses treated with steroids. Although her HLH associated symptoms rapidly responded, her EBV viraemia continued to rise. Course of rituximab successfully depleted B cells but did not reduce EBV viral load. The patient was extensively investigated for primary and secondary immunodeficiency, but no genetic causes found. Then she had 3rd HLH relapse and was commenced on etoposide as per HLH 94 protocol with resolution of clinical symptoms and reduction in EBV viral load. During the 3rd HLH relapse treatment course patient had multiple complications (renal failure requiring haemofiltration, atrial fibrillation requiring DC cardioversion, adenovirus and CMV viremias, invasive candida parapsilosis enteritis, facial palsy). She then underwent Alemtuzumab/Fludarabine/Treosulfan conditioned matched sibling donor (MSD) peripheral blood stem cell transplant in October 2014. Soon after transplant EBV reactivation that resolved after 1 dose of Rituximab (January 2015). Two years after BMT she had EBV reactivation again although at low levels (<60K/mL) and has been symptomatic with prolonged fever episode and one episode of tonsillitis. Also developed bilateral arthritis. No signs of PTLD or HLH. The patient also developed CMV reactivation in February 2015 and was given Cidofovir and Foscarnet.

### Supplementary figures

#### **Sequences description and MDS**

The majority of sequences had >90% coverage with average read depth ranging from 10x to >2000x. Sequences with an average read length less than 10x and less than 90% genome coverage were not included in further analyses, leaving a total of 76 samples to analyse. There were no differences in average read depth between groups, with the exception of tumour samples from PTLD which had, on average, a higher depth.

**Supplementary figure 1:** Boxplot showing the average depth, excluding duplicate reads and repeated regions, for each sample by clinical group.

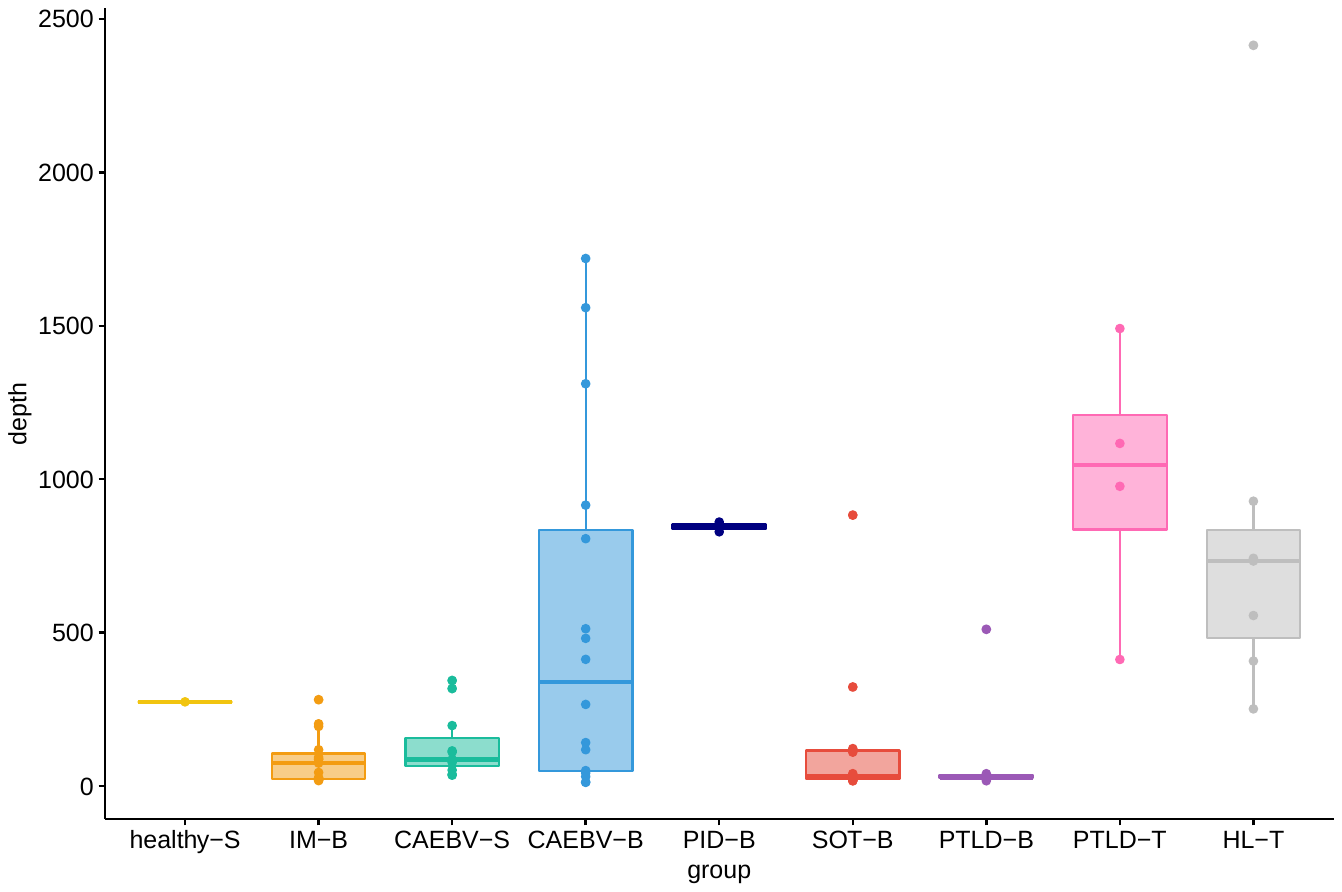

We showed that EBV genomes did not cluster by disease but rather by geographical origin. All four cases of CAEBV clustered away from the Asian genomes; the PID case although living in Europe was of Asian origin with parents who had grown up in Asia from whom the patient may have acquired the virus. One sample was found to be EBV type 2 (in the PTLD group, patient 5); this clearly clustered apart from all other viruses (data not shown) and was therefore excluded from further multidimensional clustering. Longitudinal samples from individuals, including those after hematopoietic stem cell transplant (HSCT), clustered closer together than those from unrelated patients. Sequences from one pair of saliva and blood samples from a patient with CAEBV clustered apart suggesting different consensus genome sequences.

**Supplementary figure 2**: MDS clustering of all samples coloured by patient group. Multiple samples from CAEBV patients P1, P2, P3 and the paired blood-saliva (P4) are indicated.

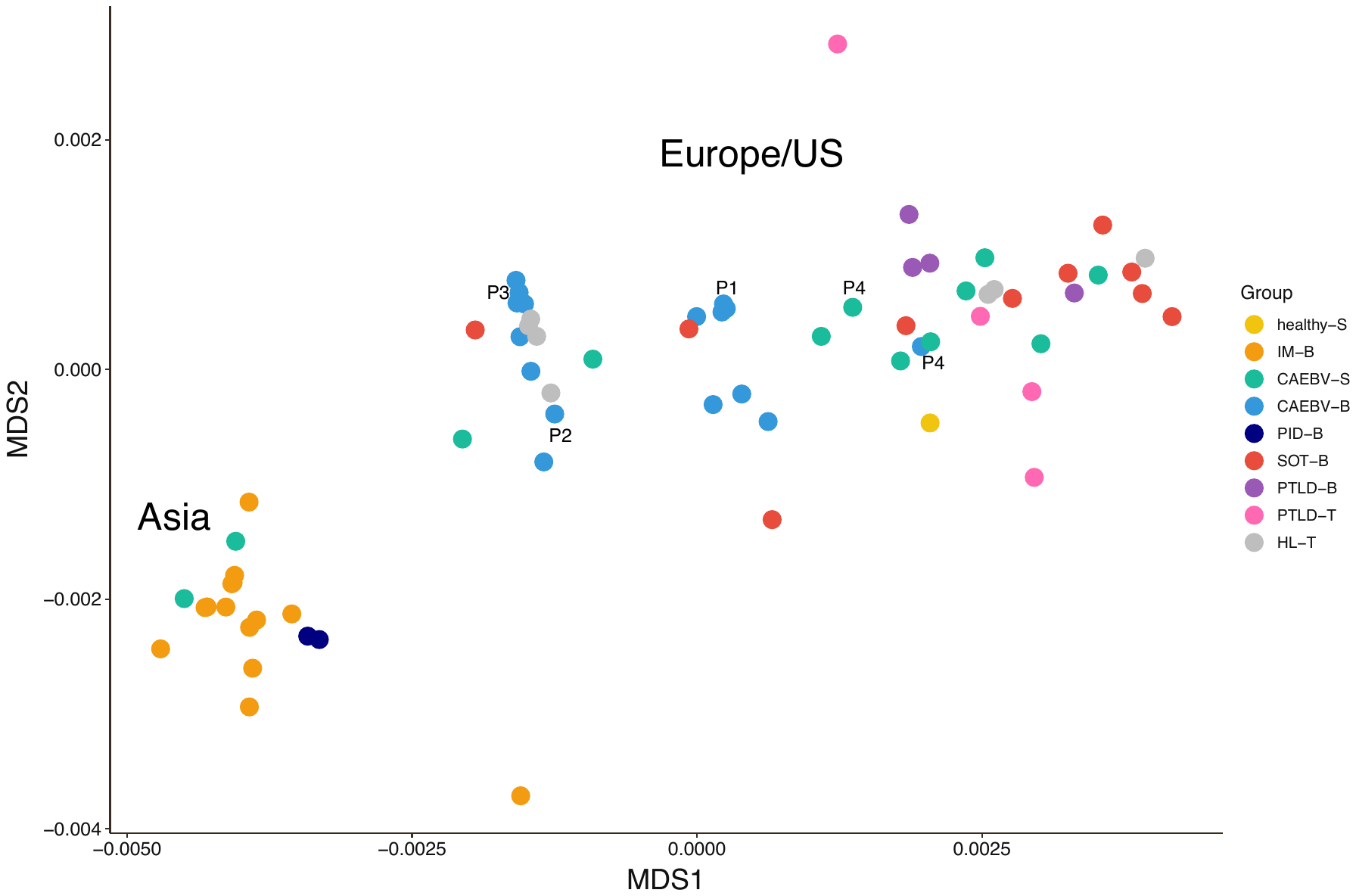

#### **Within-host diversity and mixed infections**

We have previously shown that samples with high within-host nucleotide diversity can contain mixed infections ^3^. We calculated EBV genome-wide within-host diversity (π) with a mixture model scheme which is not affected by average read depth (for details see Appendix Methods in ^3^. Excluding repetitive regions (based on GenBank sequence NC-007605.1) for which mapping anomalies tend to generate false variants, we calculated mean EBV genome-wide within-host diversity (π) to be 1.59x 10^-3^ (median=1.24x 10^-3^, SD=1.03x 10^-3^) similar to that previously observed. Overall saliva samples showed significantly more within-host nucleotide diversity (Kruskal-Wallis rank sum test, p-value < 0.001), than other samples (Figure 2). Despite having the highest read depth, tumour samples had the lowest diversity and in the case of PTLD significantly lower than for matched blood samples (Wilcox-test adjusted for multiple comparison p-value <0.05; Figure 2).

**Supplementary figure 3:** The y-axis shows the within host nucleotide diversity (π) per sample by group (x-axis and colours). Horizontal grey dashed line indicates the median diversity for all samples + 2 standard deviations (SD), which was used as threshold for investigating mixed infections. Significance bars are for Kruskal-Wallis rank sum test with FDR multi-comparison adjustment; * indicates p-values less than 0.05, ** p-values less than 0.01.

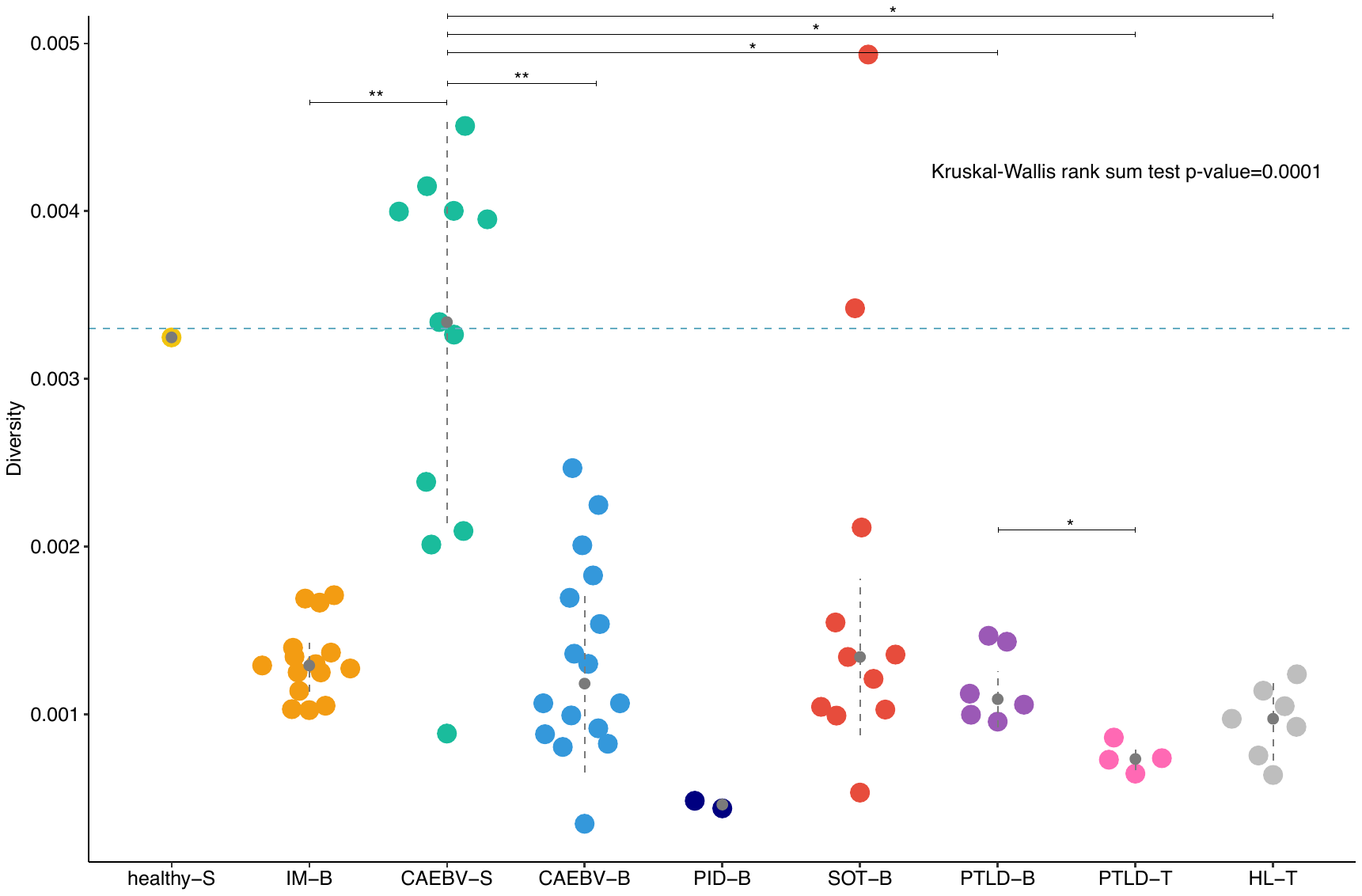

**Supplementary figure 4:** Relationship between average depth (x-axis) and diversity calculation (y-axis). Higher diversity is not associated with higher depth (minimum depth shown is 10x) (see also Supplementary table 1).

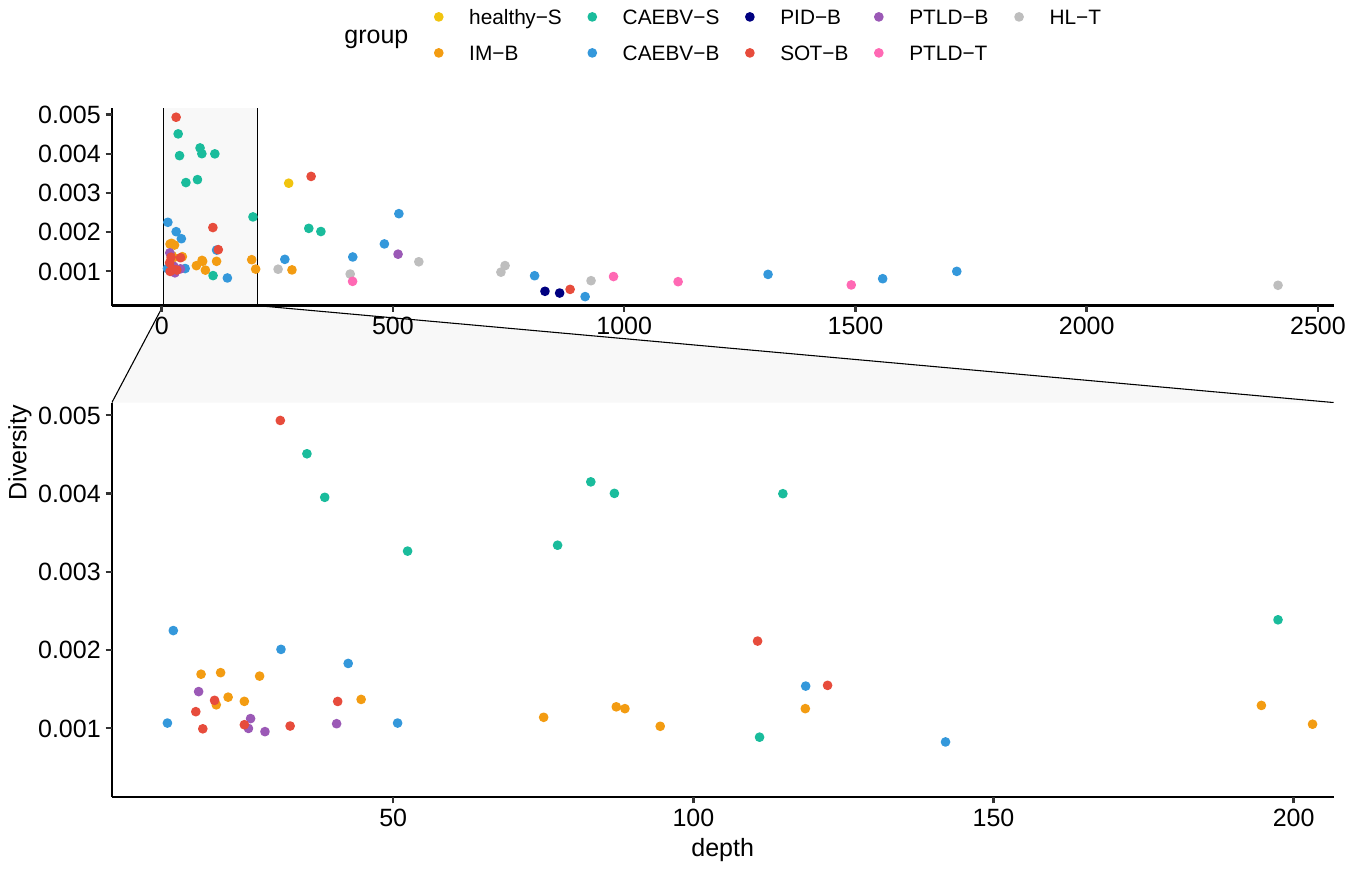

**Supplementary figure 5:** Frequency distribution of variants (obtained by mapping against their own consensus) for (A) blood SOT samples suspected as harbouring mixed infections (B) all salivary CAEBV samples and (C) saliva from an asymptomatic shedder.

A) **SOT blood samples**

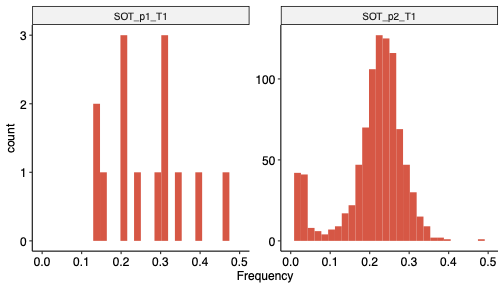

B) **CAEBV salivary samples, ordered by diversity**

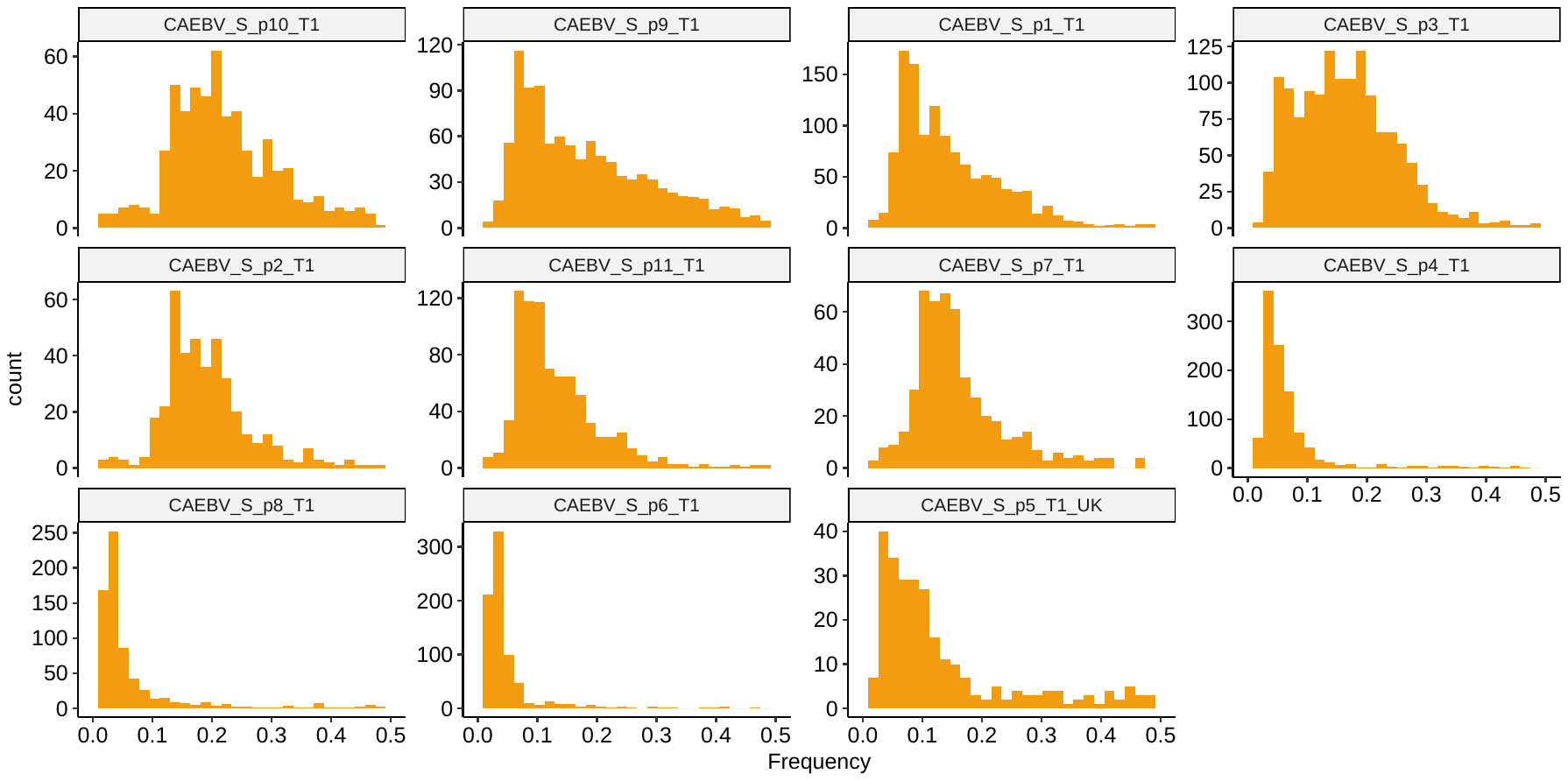

C) **Healthy salivary sample**

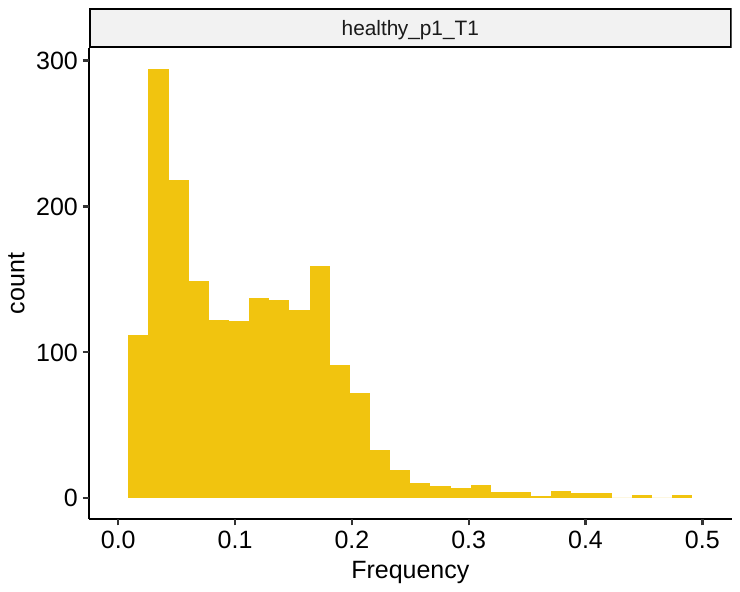

**Supplementary figure 6:** Haplotype frequency after haplotype reconstruction (HaROLD) ^3,4^ in SOT blood samples (A), saliva samples from CAEBV patients (B) and asymptomatic shedder (C). The majority haplotype is coloured pink by convention with additional haplotypes coloured green and yellow.

A) **SOT**

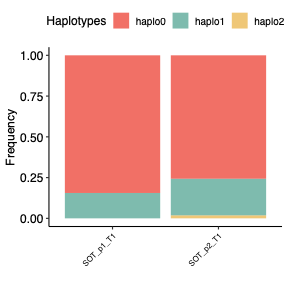

B) **CAEBV salivary samples**

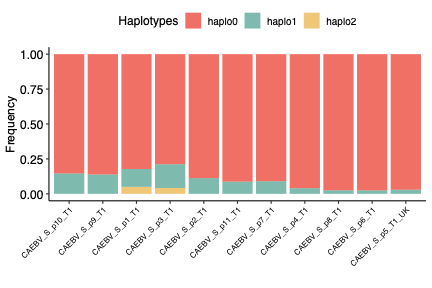

C) **Healthy salivary sample**

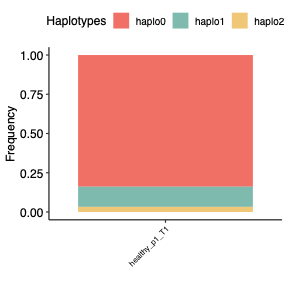

**Supplementary figure 7:** Multidimensional scaling (MDS) clustering including all EBV sequences included in the paper (in grey) and the reconstructed haplotypes sequences. Haplotypes of the same colours were derived from the same patient/sample. SOT_p1_T1 was excluded because there were not enough reads to reconstruct a good quality sequence. Where haplotypes from the same patient cluster apart this indicates the presence of distinct strains

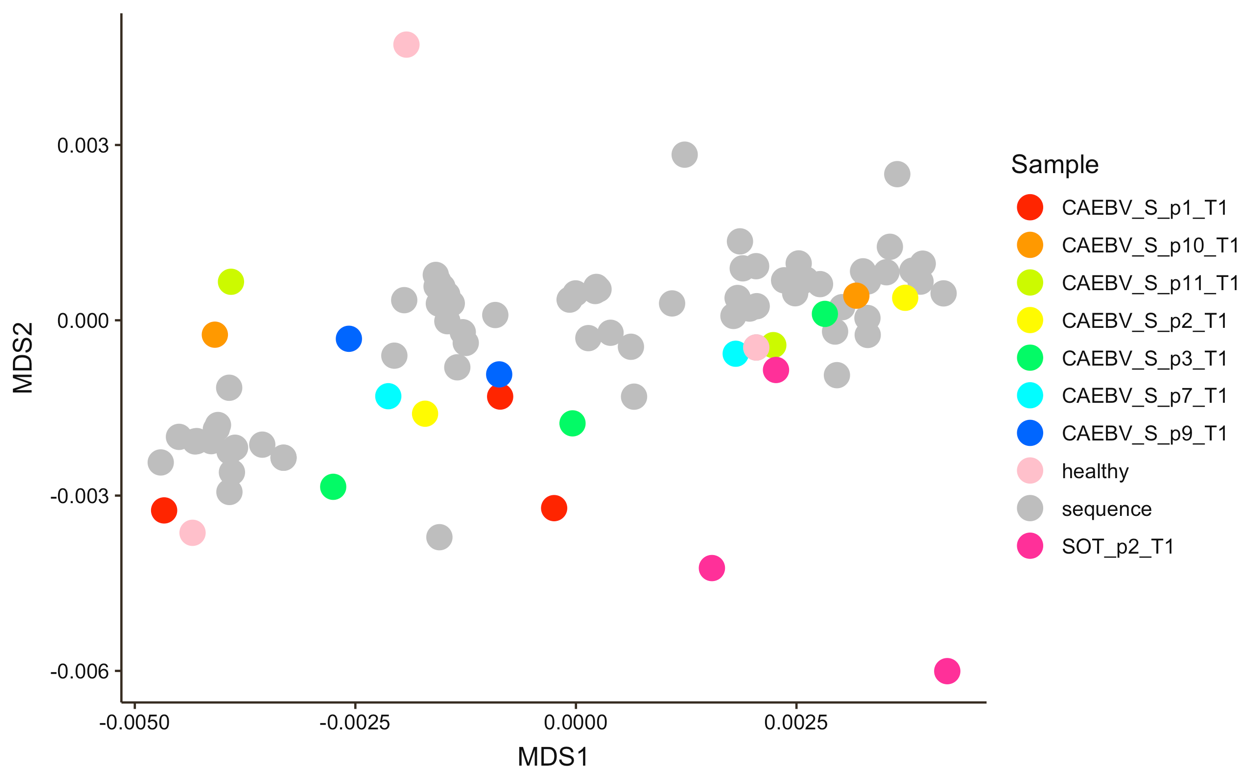

#### **Single-nucleotide variants (SNVs)**

To avoid bias due to different number of reads mapped in different samples, we only selected the samples with the highest average read depth for each patient and we plotted the relationship between the average read depth and the number of SNVs, showing that there was not obvious association. Indeed, samples with very high depth, such PTLD tumours had a very low number of SNVs and vice versa a few samples with a lower average depth in the saliva CAEBV had a greater number of mutations.

**Supplementary figure 8:** Relationship between number of SNVs in total (including all types of coding and non-coding mutations with at least 2% frequency and strict QC as explained in the Methods section) and the average depth by sample coloured by disease group (minimum average depth is 10 x)

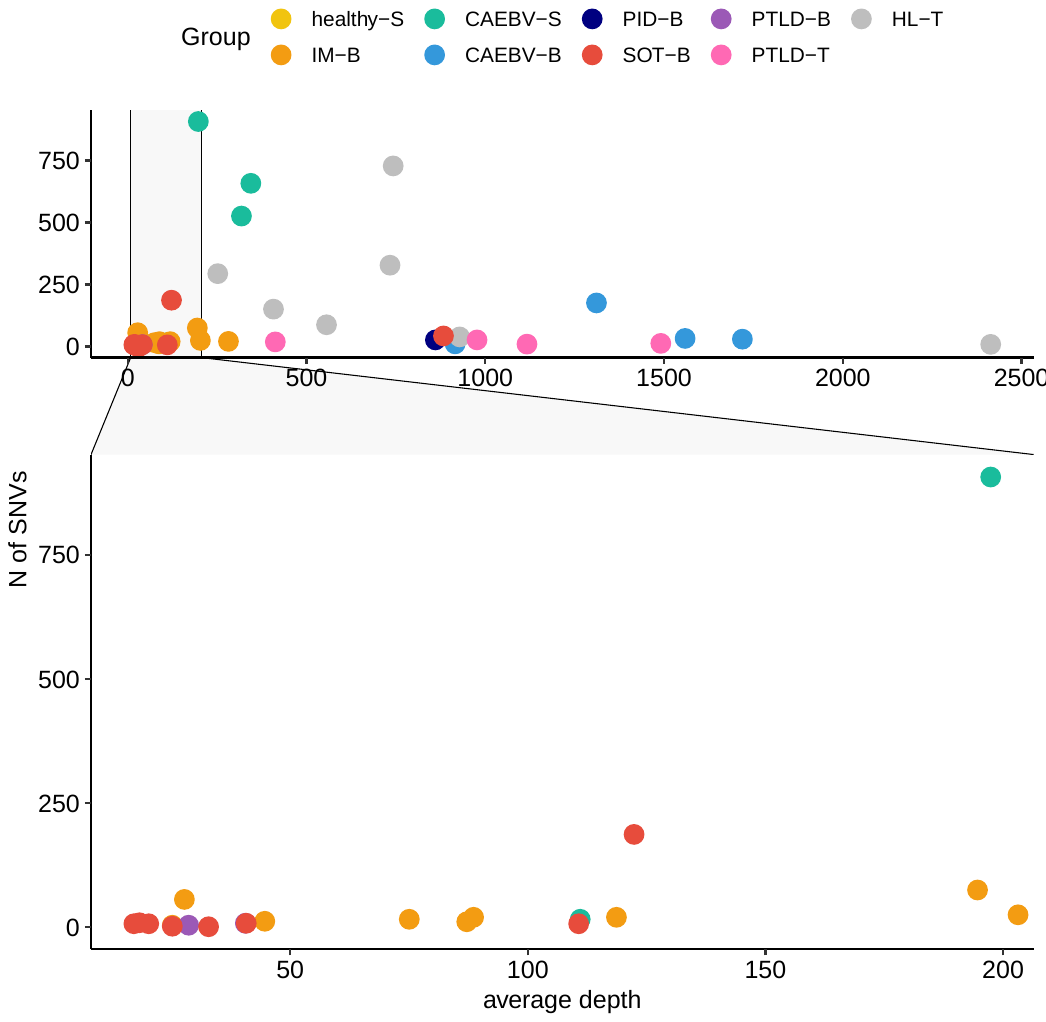

Excluding mixed infections, we detected 4633 single nucleotide variants (SNVs) from 0 to 907 per sample (mean of 113 and median 20), of which 69% affected coding sequences (17% leading to amino acid changes, according to NC_007605.1). Salivary CAEBV samples had a significantly higher number of SNVs compared to other groups (Kruskal-Wallis rank sum test), but not compared to blood samples. Likewise, HL tumour samples had a greater number of low frequencies SNVs compared to IM and SOT).

**Supplementary figure 9:** Boxplots for number of SNVs in each EBV-related disease/group. Significance: * p-value <= 0.05, ** p-value <=0.01.

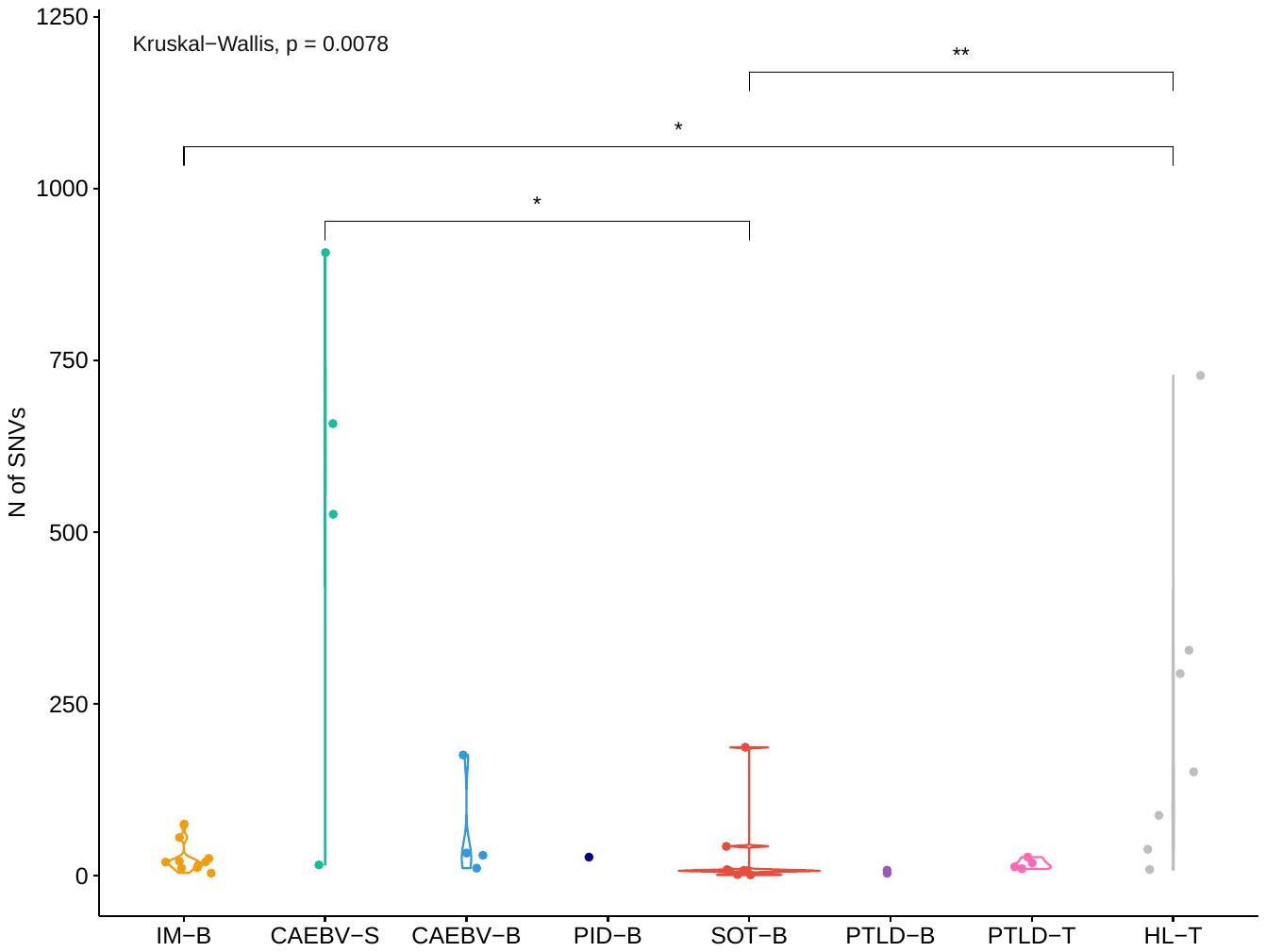

The location and nature of these SNVs in samples containing a single genotype is shown below. Non-synonymous mutations were enriched in all subject groups in latent genes, in particular EBNA3 (where the most common variants were H782P, G783R, S786T and P790Q) and BKRF1/EBNA1 (the most common amino acid substitutions were D499E and E629D). CAEBV saliva samples and HL tumours had the highest number of SNVs affecting several categories of lytic genes (tegument proteins such as BPLF1 and glycoproteins i.e. BLLF1 [gp350]). Variants in capsid proteins (including BcLF1 and BDLF1 genes) were only observed in CAEBV blood and HL tumours, although they did not have specific variants in common. Many SNVs in blood CAEBV samples were also enriched in regulatory genes, in particular BALF1.

**Supplementary figure 10:** Location of single nucleotide variants (SNVs), excluding samples with possible mixed infections in the EBV genome. Samples from infectious mononucleosis (IM), CAEBV as well as EBV positive solid organ transplant (SOT), PTLD and HL were analysed. Each line represents an EBV genome from a single patient. If multiple samples for one patient were present, only the one with the highest average depth was included. Each coloured dot indicates a variant from their own consensus sequence. Graph shows the frequency of the variants.

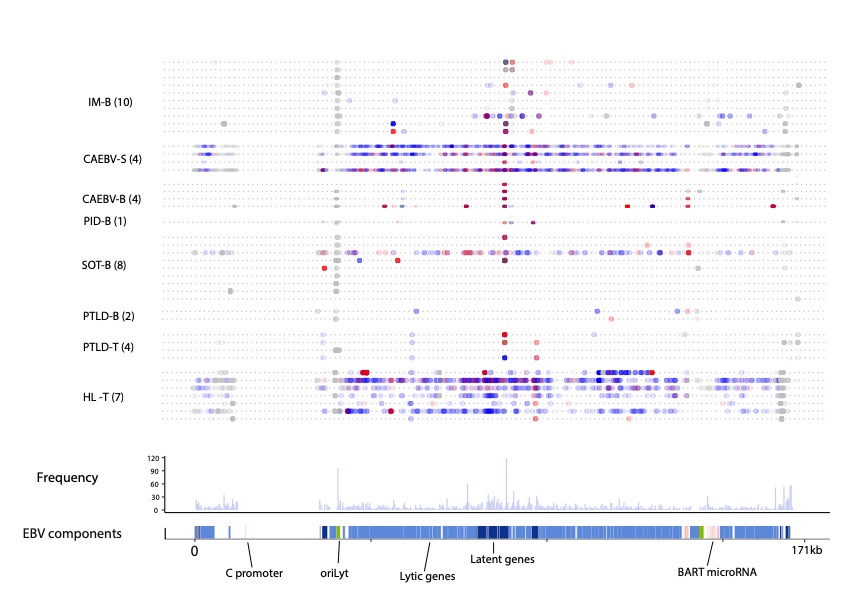

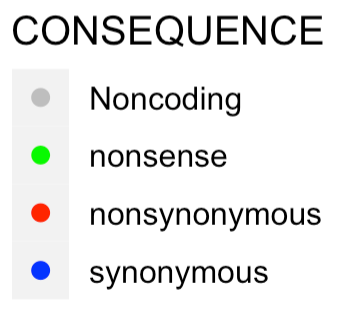

**Supplementary figure 11:** The most common non-synonymous mutations are coloured by disease group.

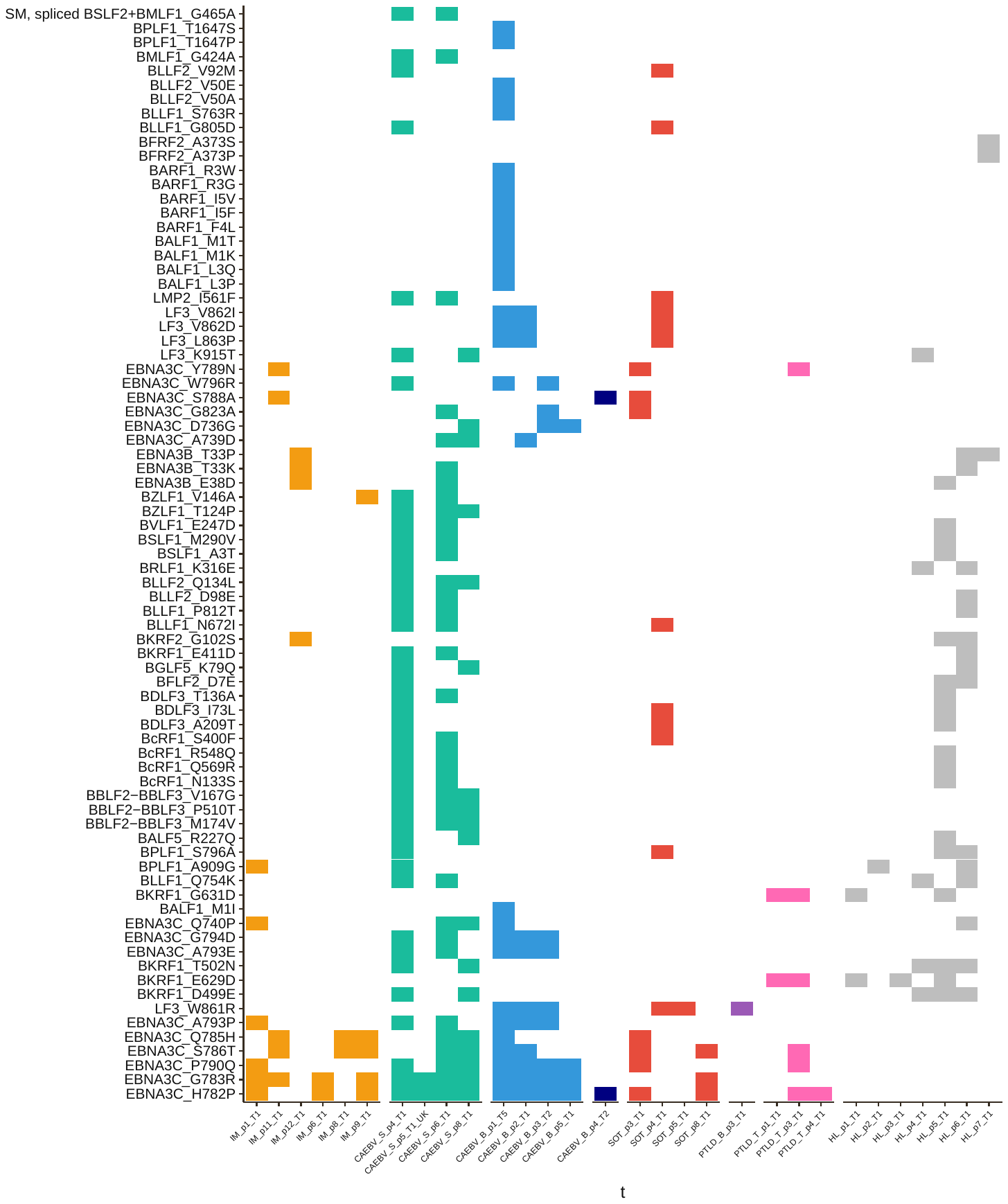

**Supplementary figure 12:** Total number of non-synonymous SNVs by sample (only samples with non-synonymous variants are shown) and genes involved by category. The size of the filled circles represents the number of variants.

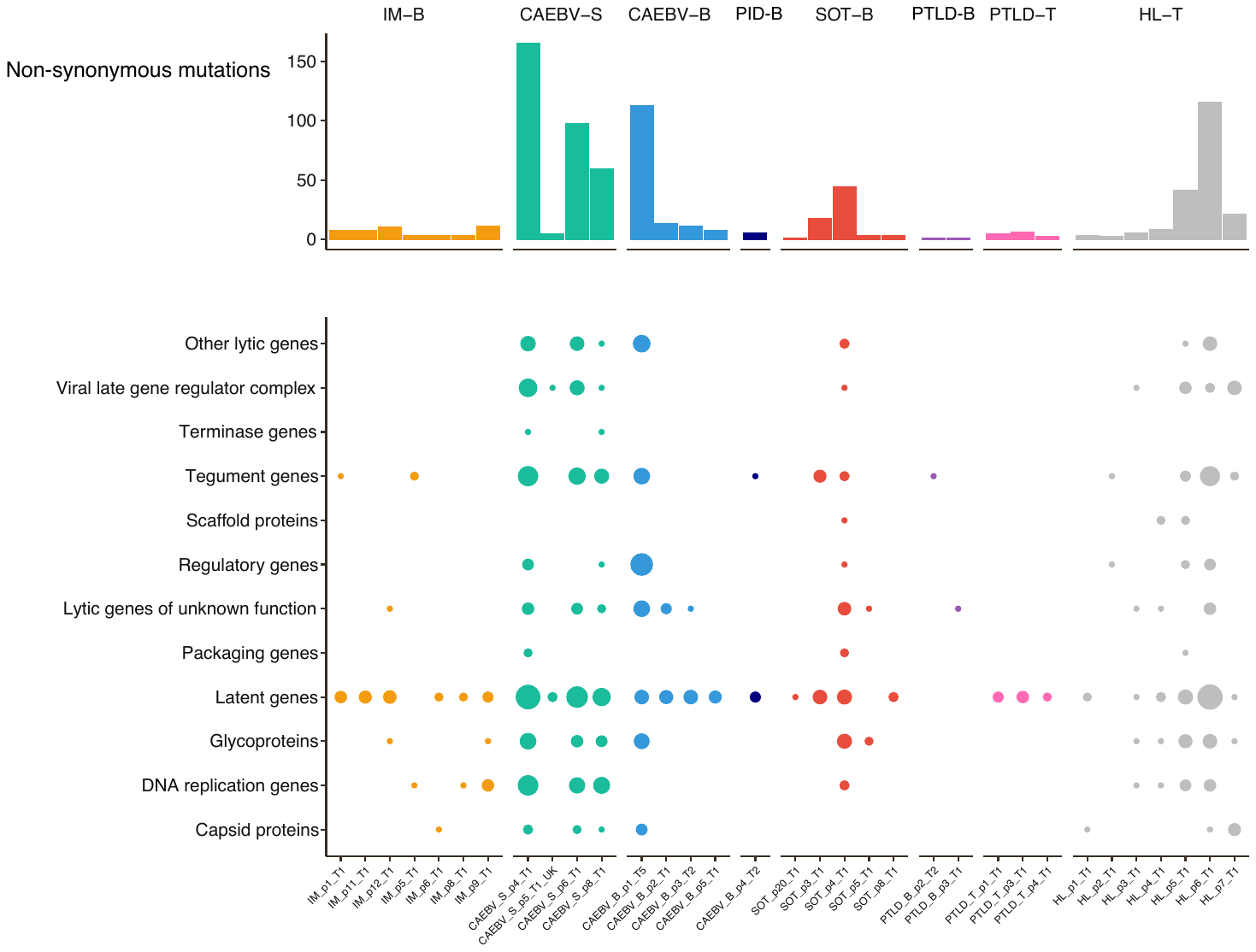

#### **Deletions**

Deletions of between 1 and 30 bp (indels) occurred at similar frequencies in all samples, suggesting that these were method-related artefacts, and they were thus excluded from subsequent analysis (Supplementary figure 14). A median of 9 deletions ≥ 30bp was observed per EBV genome with most seen in PTLD tumours (median 38.5), with the fewest seen in PTLD and SOT blood samples (median respectively of 1.5 and 4.5), including those paired with tumour samples.

Deletions were subdivided into those ≤2kb in length and greater than >2kb in length based on the size distribution (Supplementary figure 15). All EBV genome deletions of ≤2 kb were mostly in-frame and clustered in the same latent and structural gene hotspots as seen for SNVs. This included a 30 bp deletion in LMP1, primarily found in Asian strains, which we detected, predominantly at the consensus level, in 58% of the samples: 70% of Asian samples have this deletion versus 22% of European samples. We also identified a sporadic deletion in CAEBV which affects glycoprotein H (gp85 BXLF2) (66 bp in CAEBV patient 1).

Deletions >2 kb were predominantly present at low frequencies in malignancies and CAEBV, including PTLD tumour-tissue (4/4), HL tissue (4/7) and CAEBV blood (5/5). One deletion located within an intergenic region 12118-15159, within the major internal repeat, was shared by EBV sequenced from blood of all five CAEBV patients, but no other samples, including CAEBV saliva samples (Supplementary figure 15). Large deletions were identified within the region 120470 to 158062, which codes for several lytic genes, including scaffold proteins (i.e. BdRF1, BVRF2), glycoproteins (i.e. BILF2, BXLF2), a tegument protein (BVRF1), and regulators of late gene transcription (BcRF1, BVLF1). Although the deletions varied in size, these EBV genes were affected in samples from CAEBV blood (1/5), PTLD tumour-tissue (2/4) and HL tumour-tissue (1/7). This region also includes the BART miRNA clusters which were often deleted in tumour samples (both PTLD and HL), but also in CAEBV blood (1/5).

**Supplementary figure 13:**

Size of deletions in all samples, coloured by group. First plot (at the top) shows the full range of deletion’ sizes (1-24474). The second plot is a zoom for deletions with sizes between 1 and 100. Indels with a size smaller than 30bp were present in all groups and considered artefacts.

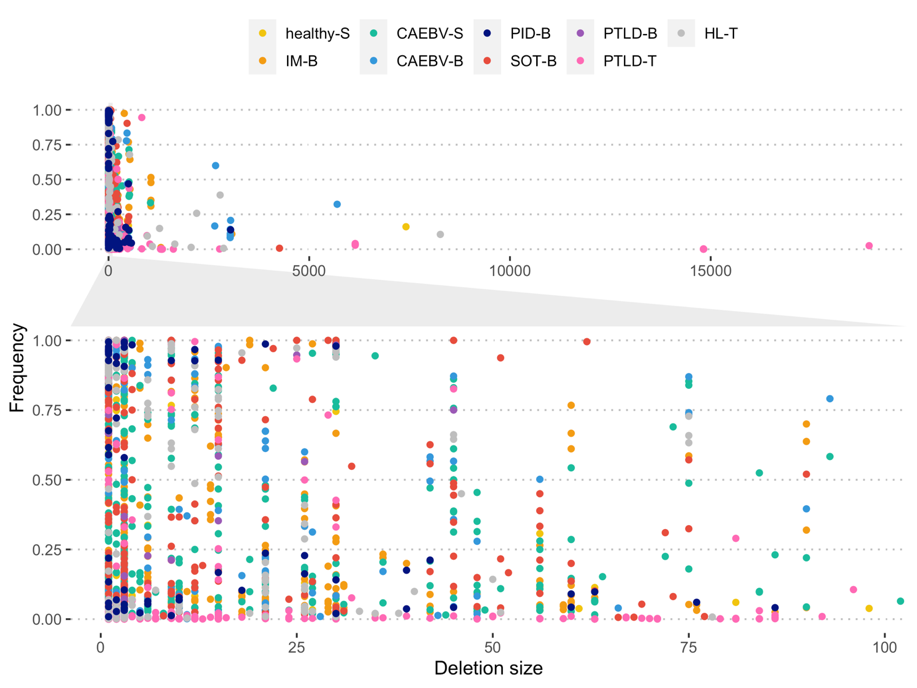

**Supplementary figure 14:** Distribution of size of deletions (left – all deletions, right – a zoom of sizes from 30 to 10k bp). Most (95%) deletions were smaller than 2k bp, therefore that was chosen as cut-off to distinguish “small” deletions vs “big” deletions.

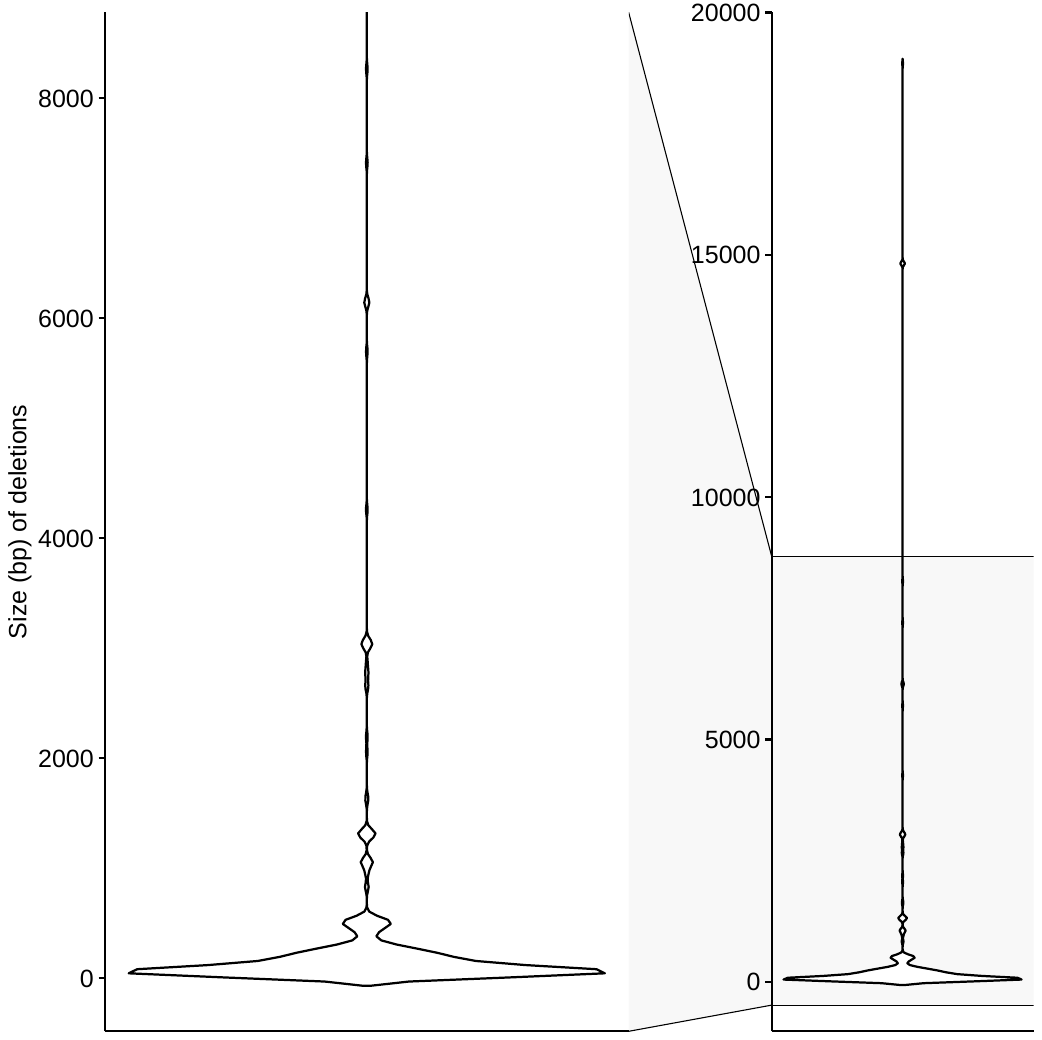

**Supplementary figure 15:** Number of triplets (x-axis) vs number of non-triplets (y-axis) by gene and coloured by group.

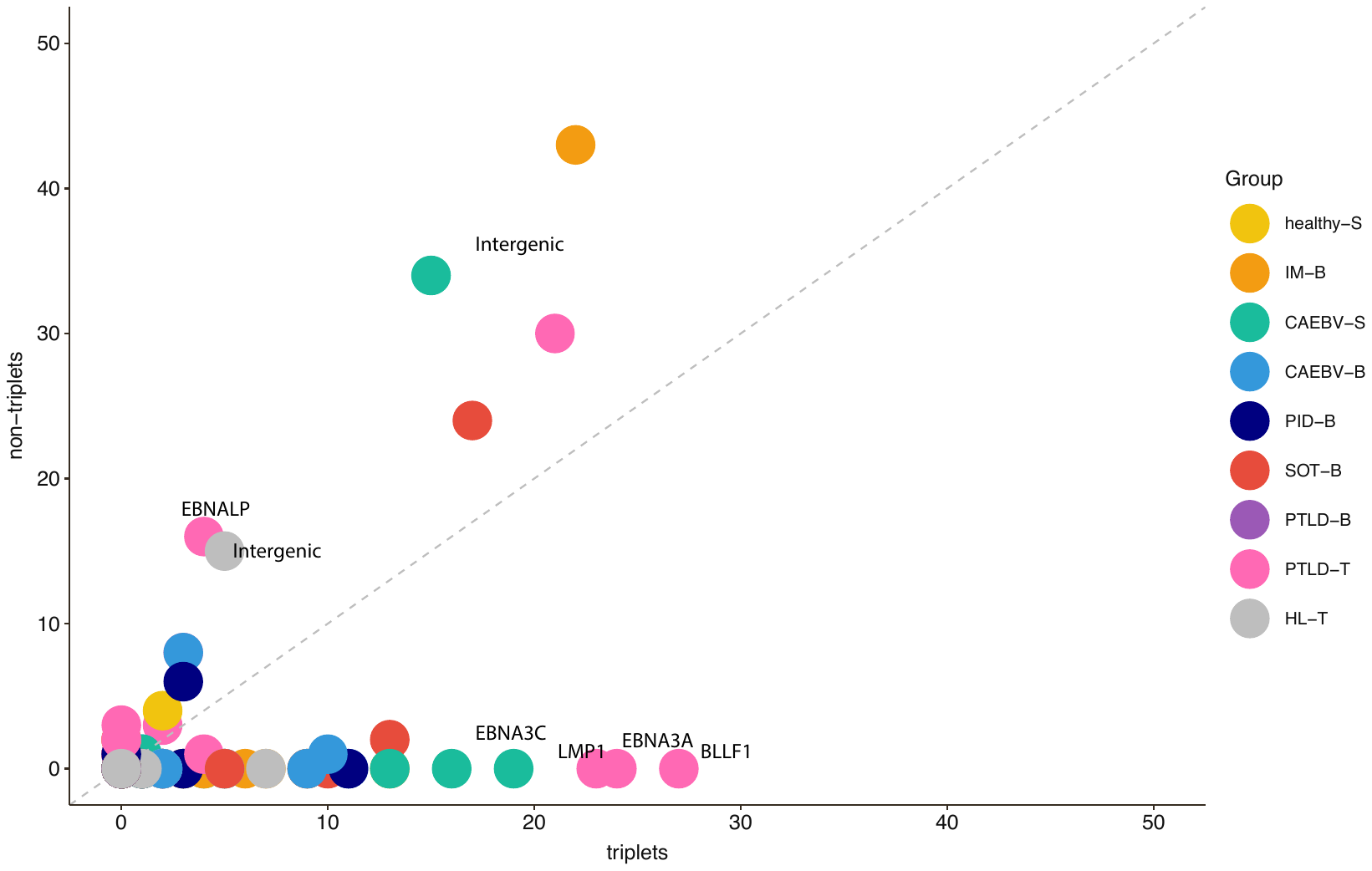

**Supplementary figure 16:** Total number of deletions (>=30 bp and < 2k bp in size) per sample coloured by clinical group (A) and the genes affected by deletions (B).

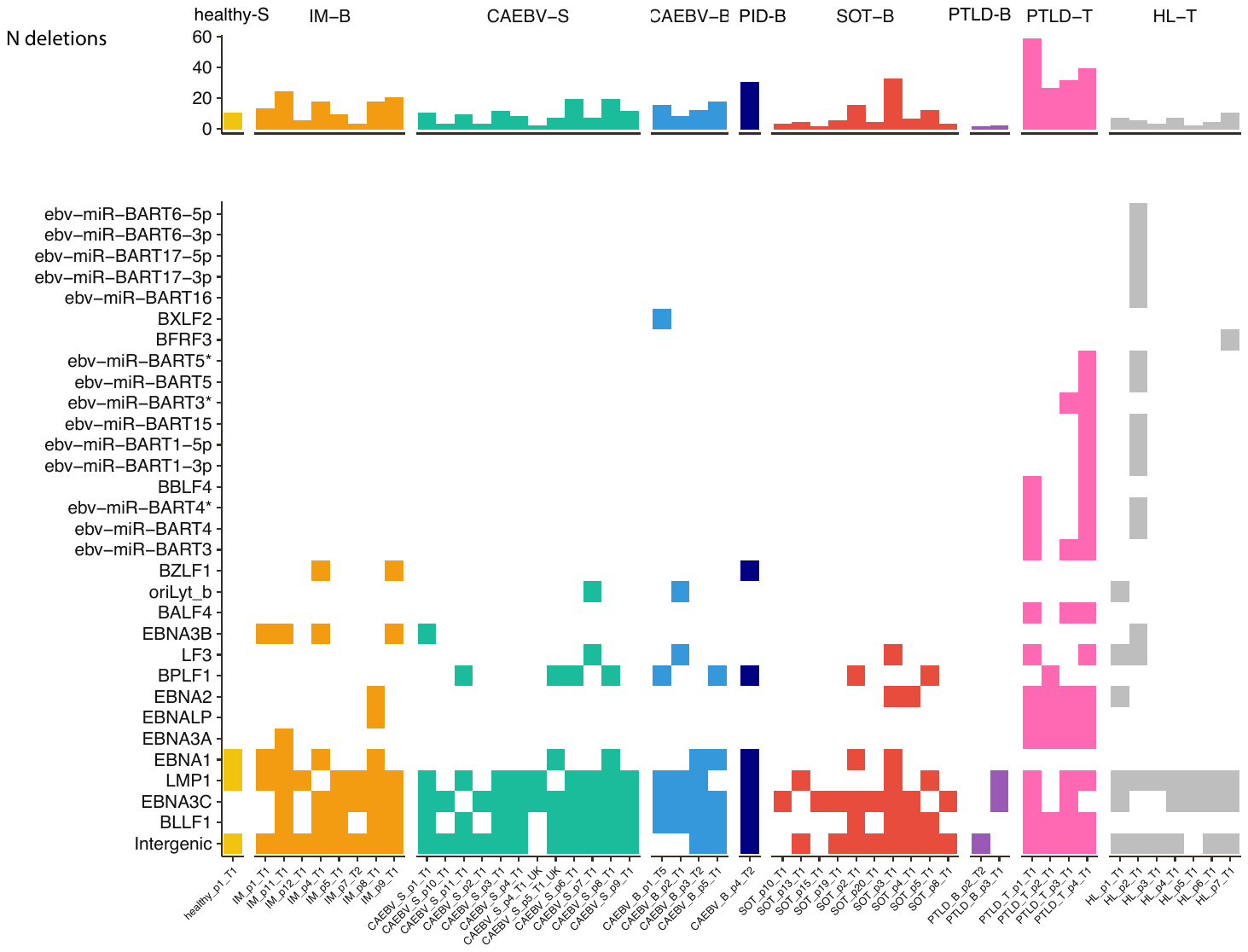

**Supplementary figure 17:** Genes affected by larger deletions (> 2k bp) in each sample coloured by clinical group.

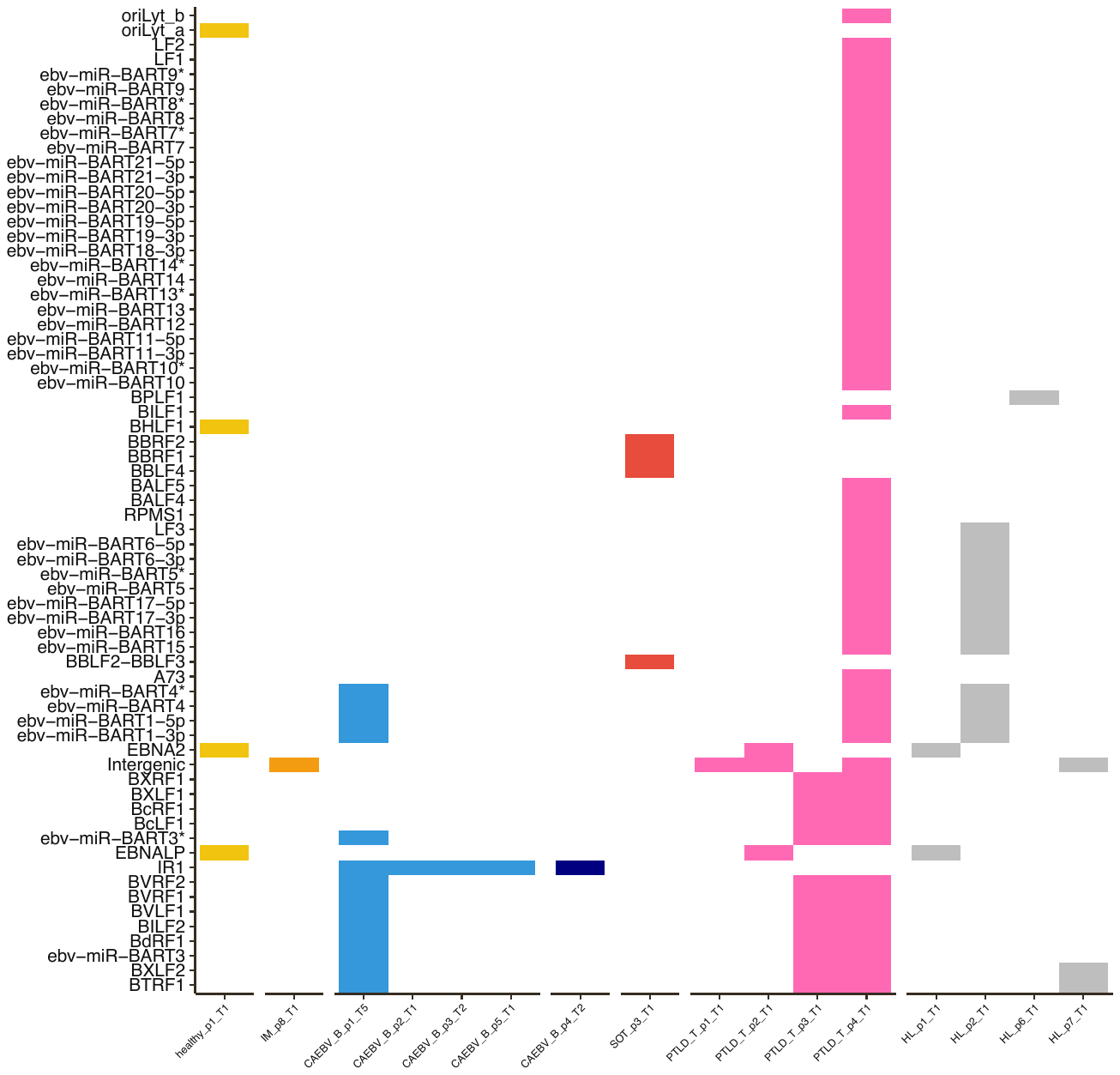

**Longitudinal data**

**Supplementary figure 18:** Analysis of longitudinal samples in CAEBV patients. Samples from three patients are shown in different colours. The top panel (A) shows the number of deletions for each sample, including all deletions larger than 30 bp. Genes affected by larger deletions (> 2k bp) are shown in panel B; over time all larger deletions are lost. SNVs are shown in (C) and (D).

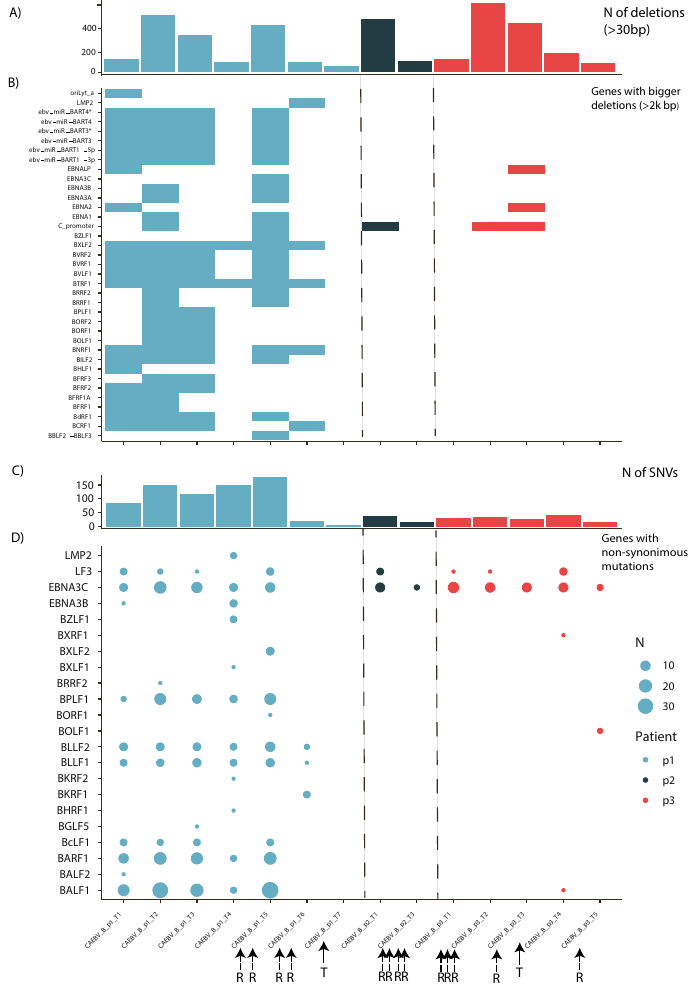

**Supplementary figure 19A**: Number of total SNVs and genes affected by non-synonymous mutations for longitudinal blood samples for PTLD and IM patients.

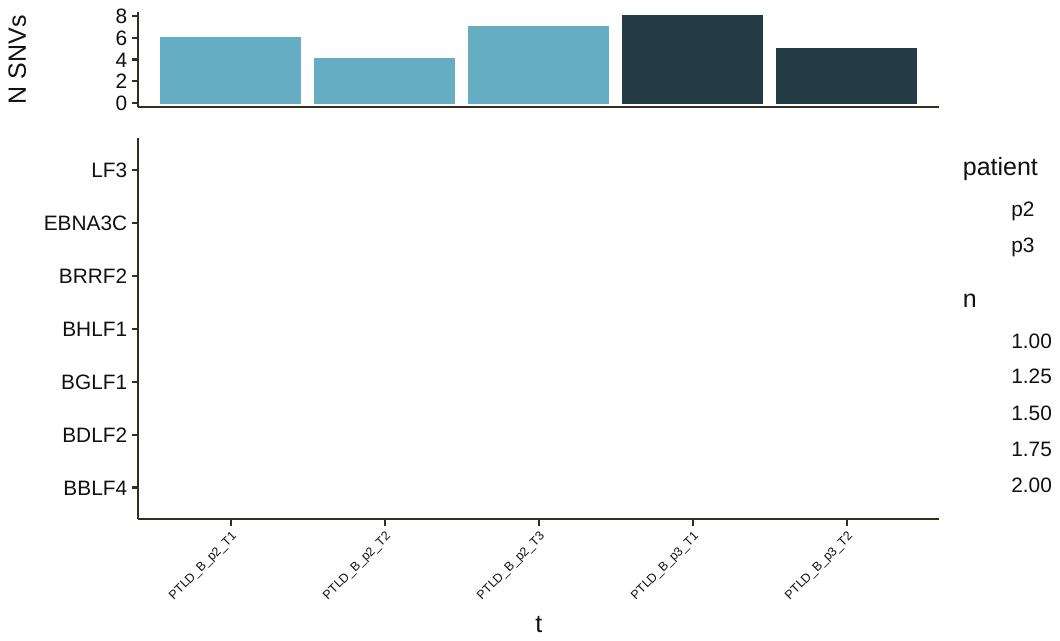

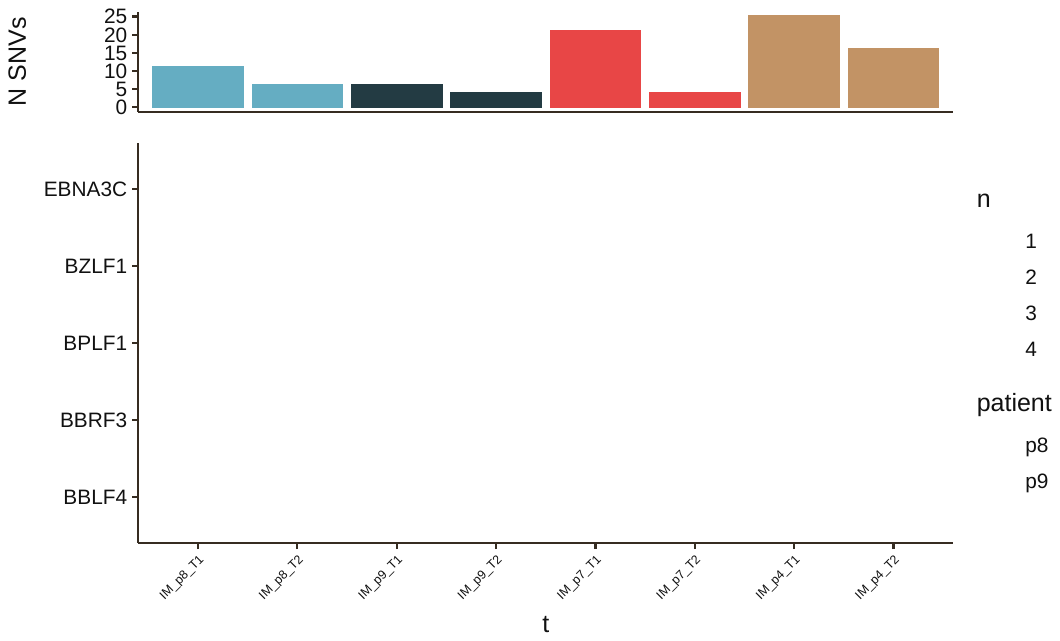

**Supplementary figure 19B**: Total number of bigger deletions (>2k bp) and genes affected by deletions in longitudinal blood samples for IM and PTLD patients.

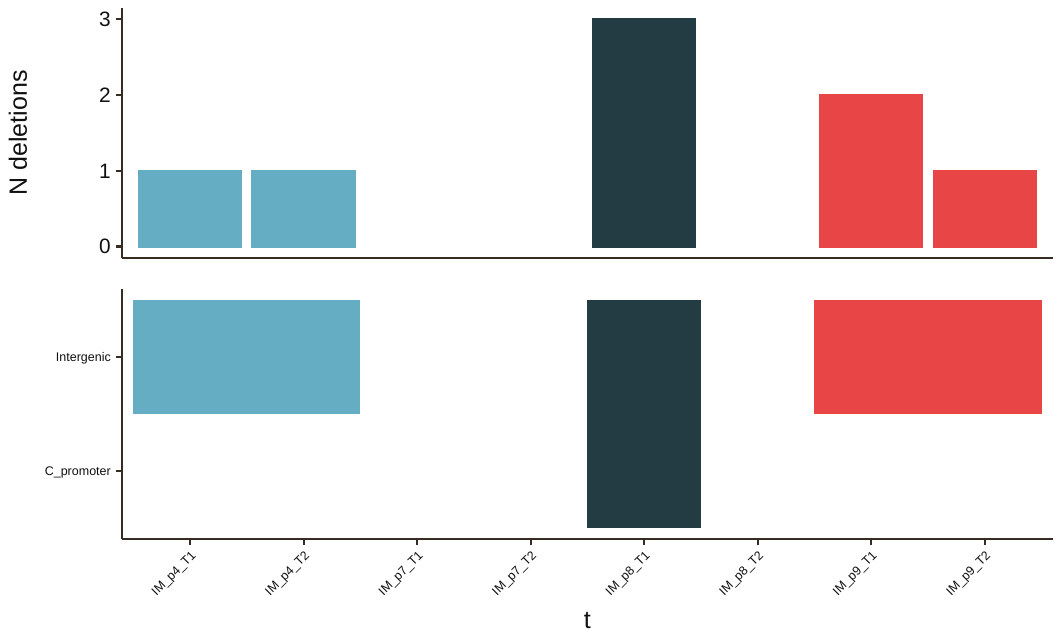

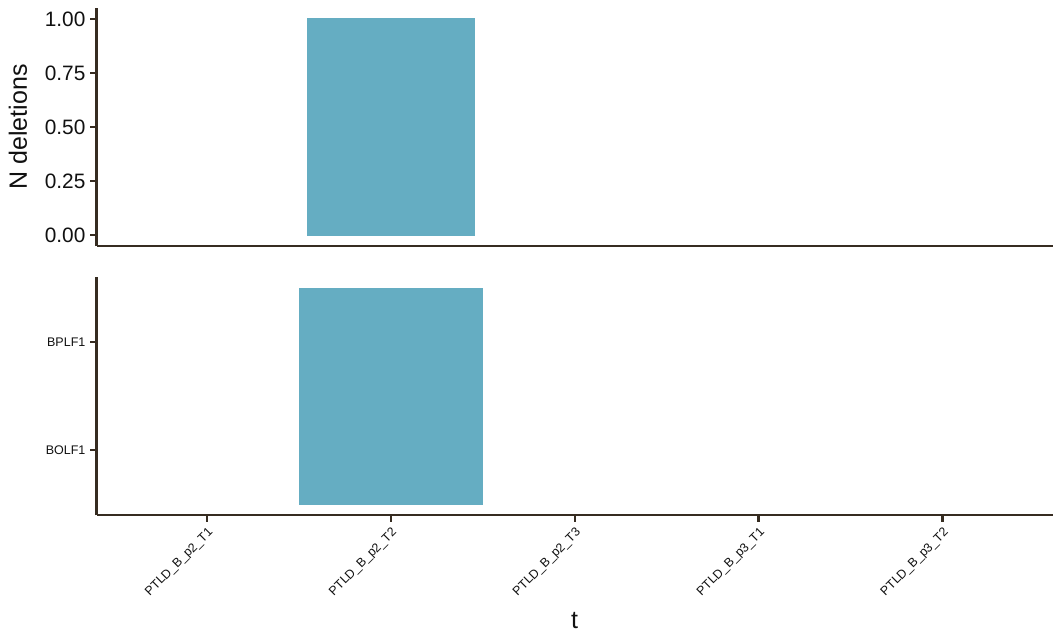

**EBV in different compartments**

We compared genomic data from whole blood, T cells and saliva in one patient with PID-B and whole blood and saliva in CAEBV patient 4.

In the PID-B patient, both whole blood and T cell separated samples had a similar average read depth of >800x. The consensus sequences were almost identical, with only four variants of which three were in repeat regions. Minority variants were similar in both samples with some differences occurring only at a very low level, especially in the area 88360-88523 bp (overlapping with gene EBNA3C). We found virtually identical deletions in both samples (in total: 89 for whole blood sample and 82 for T cells). Most deletions were present at similar frequencies, including a 30 bp deletion in LMP1 at higher frequency (>90%) which is common in Asian sequences. In addition, larger deletions including the 12118-15159 which we identified in all blood CAEBV sequences, were present at low level in both whole blood and T cells. A large deletion (32285-35357), which overlaps with a non-coding region of the EBV genome present in whole blood could not be assessed in the T cell sample because of poor sequence quality in that region. Although the sequence from a matched saliva sample was of poor quality, we noted 97 SNV differences compared to consensus sequence in blood, mostly in the repeat regions which are problematic to assemble. We also observed the same consensus LMP1 30 bp deletion that was present in both blood samples, in the saliva sample, suggesting that the consensus sequence is the same in blood and saliva samples.

We also compared saliva and whole blood samples from CAEBV patient 4. Both samples had good average read depth (saliva sample 111x, blood 1915x) and the consensus sequences showed 46 well-supported differences, mostly in repeated regions, but also in the gene LF3. The saliva sample had a total of 61 deletions compared to 69 in the whole blood sample. The latter deletion showed evidence for the 12118-15159, found in all blood samples with CAEBV; no deletions >2 kb was found in saliva.

**Supplementary figure 20:** Summary of total number of deletions (A) and their position in the EBV genome (B) for CAEBV patient 4 in whole blood and saliva.

A)

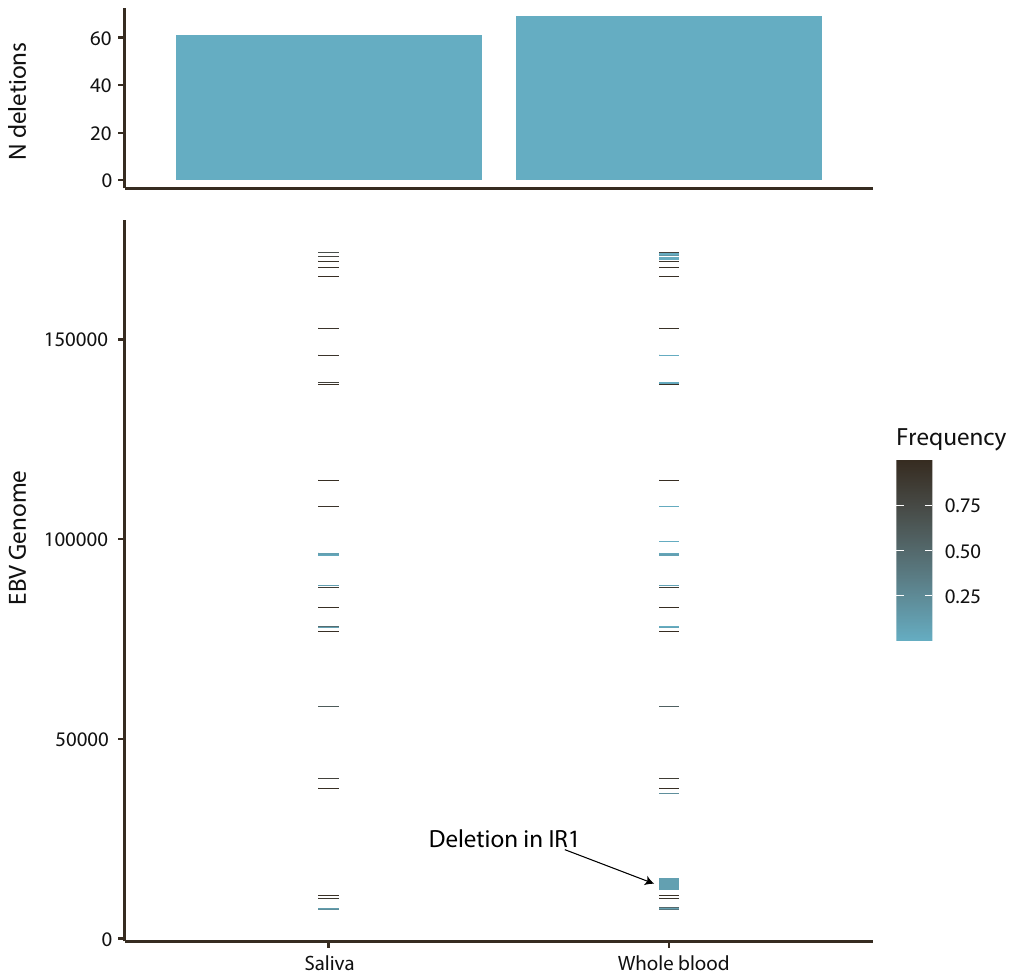

B)

**Supplementary figure 21:** Summary of total number of deletions (A) and their position in the EBV genome (B) for the PID-B patient in whole blood, T cells and saliva.

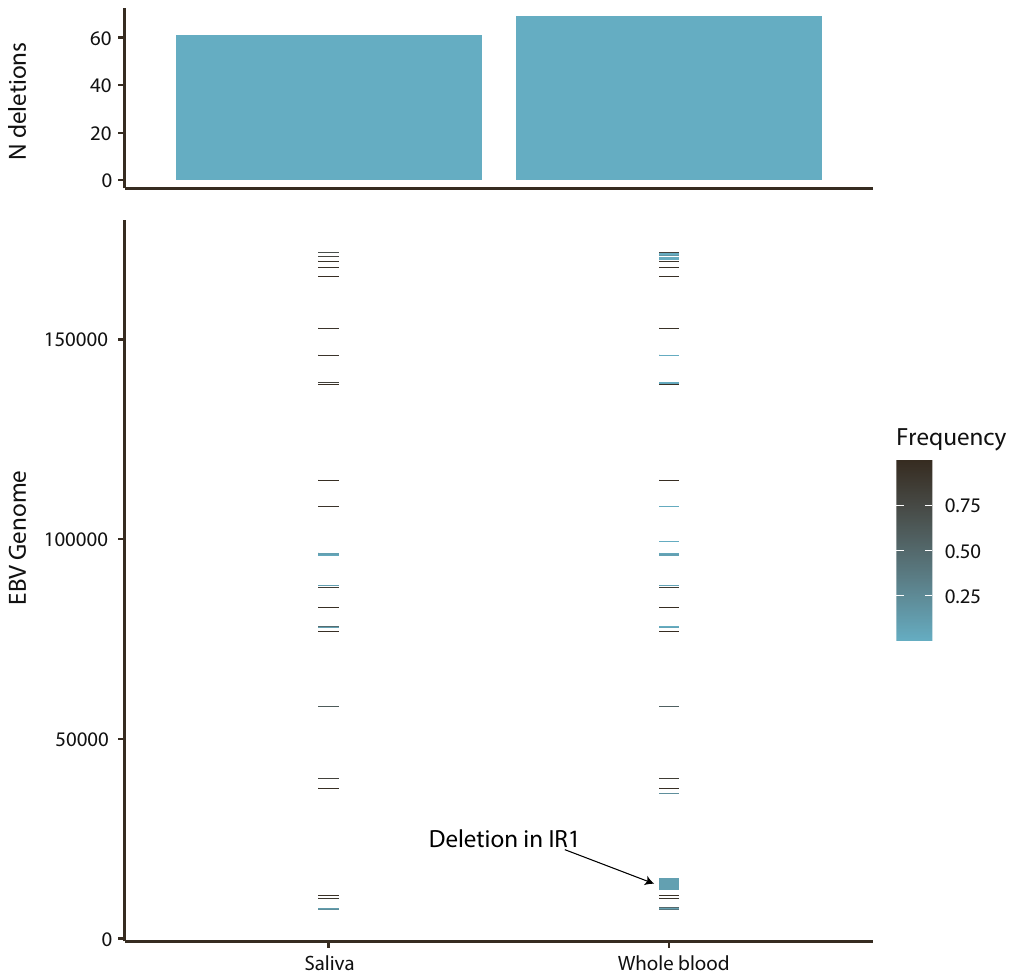

A)

B)

**Supplementary figure 22:** SNVs in whole blood and T cells in the PID-B patient. Y-axis shows the position in the EBV genome (NC_007605.1) and the colour the type of mutation. The transparency indicates the frequency.

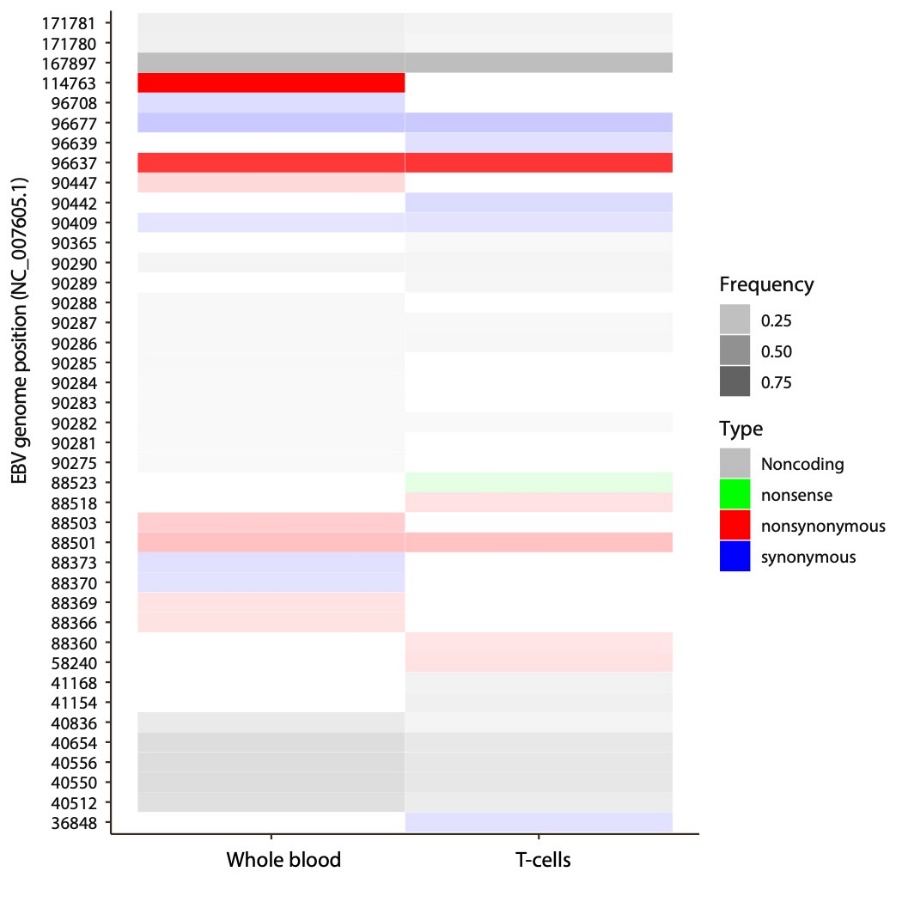
